# mRNA aggregates harness danger response for potent cancer immunotherapy

**DOI:** 10.1101/2023.03.12.23287108

**Authors:** Hector R. Mendez-Gomez, Anna DeVries, Paul Castillo, Brian D. Stover, Sadeem Qdaisat, Christina Von Roemeling, Elizabeth Ogando-Rivas, Frances Weidert, James McGuiness, Dingpeng Zhang, Michael C. Chung, Derek Li, Chong Zhang, Christiano Marconi, Yodarlynis Campaneria, Jonathan Chardon-Robles, Adam Grippin, Aida Karachi, Nagheme Thomas, Jianping Huang, Rowan Milner, Bikash Sahay, W. Gregory Sawyer, John A. Ligon, Natalie Silver, Eugenio Simon, Brian Cleaver, Kristine Wynne, Marcia Hodik, Anette Molinaro, Juan Guan, Patrick Kellish, Andria Doty, Ji-Hyun Lee, Sheila Carrera-Justiz, Maryam Rahman, Sebastian Gatica, Sabine Mueller, Michael Prados, Ashley Ghiaseddin, Duane A. Mitchell, Elias J. Sayour

## Abstract

Messenger RNA (mRNA) has emerged as a remarkable tool for COVID-19 prevention but its use for induction of therapeutic cancer immunotherapy remains limited by poor antigenicity and a regulatory tumor microenvironment (TME). Herein, we develop a facile approach for substantially enhancing immunogenicity of tumor-derived mRNA in lipid-particle (LP) delivery systems. By using mRNA as a molecular bridge with ultrapure liposomes and foregoing helper lipids, we promote the formation of ‘onion-like’ multi-lamellar RNA-LP aggregates (LPA). Intravenous administration of RNA-LPAs mimics infectious emboli and elicits massive DC/T cell mobilization into lymphoid tissues provoking cancer immunogenicity and mediating rejection of both early and late-stage murine tumor models. Unlike current mRNA vaccine designs that rely on payload packaging into nanoparticle cores for toll-like receptor engagement, RNA-LPAs stimulate intracellular pathogen recognition receptors (RIG-I) and reprogram the TME thus enabling therapeutic T cell activity. RNA-LPAs were safe in acute/chronic murine GLP toxicology studies and immunologically active in client-owned canines with terminal gliomas. In an early phase first-in-human trial for patients with glioblastoma, we show that RNA-LPAs encoding for tumor-associated antigens elicit rapid induction of pro-inflammatory cytokines, mobilization/activation of monocytes and lymphocytes, and expansion of antigen-specific T cell immunity. These data support the use of RNA-LPAs as novel tools to elicit and sustain immune responses against poorly immunogenic tumors.

---

Delivery of messenger RNA (mRNA) in lipid-nanoparticles (LNP) to antigen presenting cells (APCs) is dependent on localization to dendritic cells (DCs) and endosomal release to mediate efficient gene expression.^1-3^ Typically these LNP designs contain: 1) neutral or anionic surface charges to make particles inert at physiologic pH; 2) cholesterol/polyethylene glycol to prevent particle aggregation and maintain LNP size (∼200 nm) for endocytic uptake, and 3) helper lipids to prevent mRNA endo-lysosomal degradation.^4,5^ While this approach is effective in maximizing SARS-CoV-2 full-length spike mRNA expression to prevent COVID-19, the same approach has significant limitations for therapeutic cancer immunotherapy.^6^ Unlike the current COVID-19 mRNA vaccines, which are optimized for maximal gene expression and minimal innate immune stimulation, cancer antigens are already expressed and intertwined with a tolerogenic immune system.^6^ Using systemically (i.v.) administered cationic particles, we have sought to overcome these barriers.^7,8^ Previously, we manufactured RNA-nanoparticles (100-200 nm, +27 mEV) to elicit DC transfection, antigen-specific T cell induction and long-term survivor benefit in preclinical models.^9,10^ Our and others’ approach remains limited by the amount of mRNA payload that can be packaged per particle, which can limit antigenicity. Limits on packaging are necessary to retain nanoparticle size which is important for endosomal uptake and toll-like receptor (TLR) activation. Herein, we develop a facile approach for substantially enhancing immunogenicity of tumor derived mRNA in lipid-particle (LP) delivery systems. By using mRNA as a molecular bridge with cationic liposomes and foregoing helper lipids, cholesterol, and polyethylene glycol (PEG), we promote the formation of significantly increased self-assembling multi-lamellar RNA lipid particle aggregates (LPAs) (**Fig. 1a**). These ‘onion’ shaped particles form 200-500 nm aggregates (**Fig. 1a-b, Extended Data Fig. 1a**), with zeta potentials of ∼50 meV) (**Fig. 1c, Extended Data Fig. 1b**). By aggregating, particle size increases, and all mRNA (even Cas9-sgRNA, and plasmid DNA) were successfully loaded into multi-lamellar ‘onion-like’ vesicles (**Fig. 1d**). This design overcomes a key barrier in nanotherapeutic delivery and enhances the amount of payload that can be feasibly loaded.

**Fig. 1:**
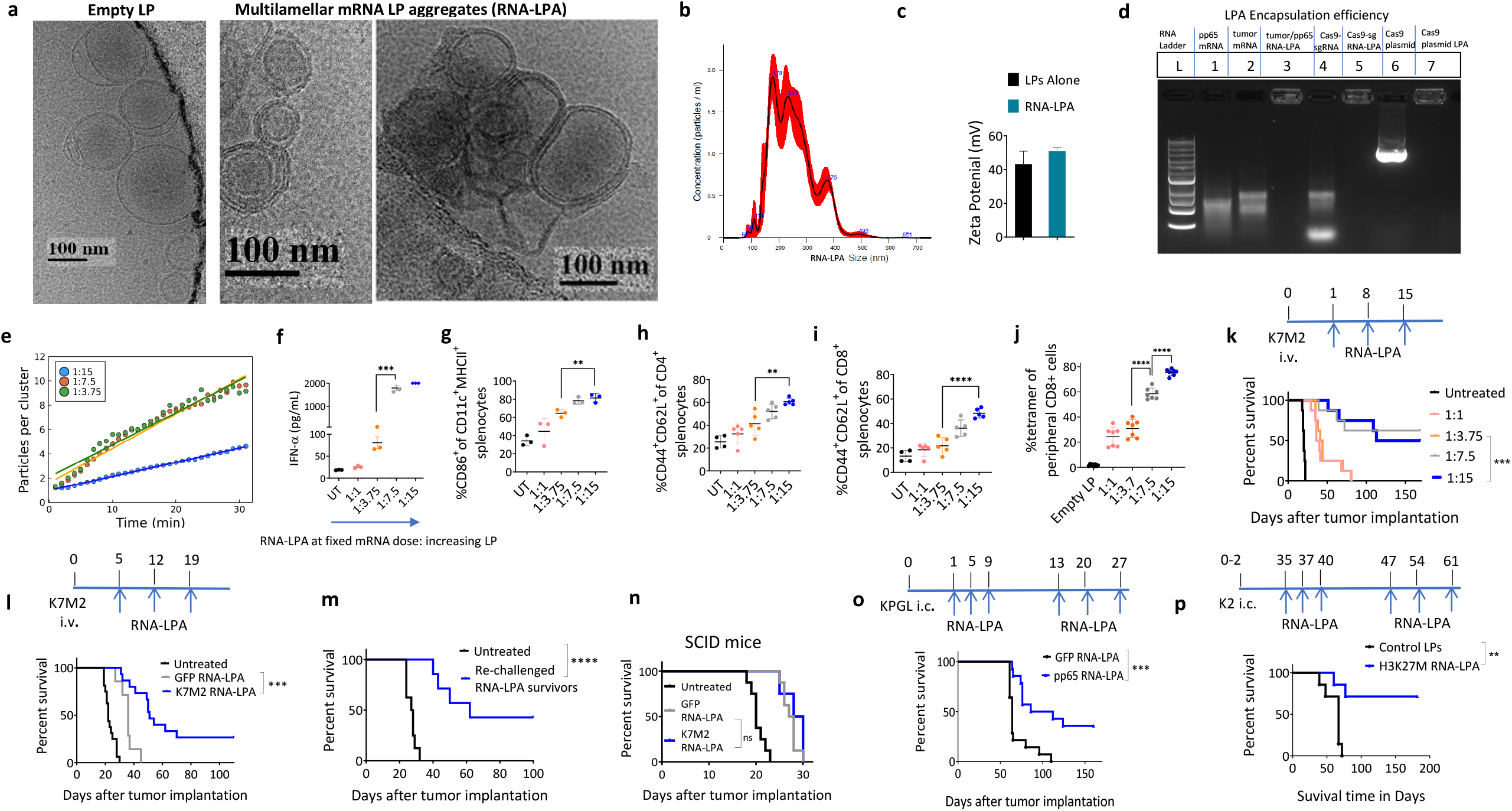
RNA-LPAs optimize payload packaging for induction of anti-tumor immunity across preclinical cancer models. **a**, Cryo-electron micrographs. **b**, nanosight size distributions. **c**, zeta potential measurements. **d**, encapsulation efficiency of RNA-LPA at 1:15 mass ratios. **e**, RNA-LPA cluster growth over time at increasing LP:RNA ratios. **f**, IFN-α measurements from serum of Balb/c mice (n= 3) within 6h of i.v. RNA-LPA loaded with total tumor derived mRNA from K7M2 (upper limit of detection for assay is 2,000 pg/mL). **g**, Activated DCs. **h**, central memory CD4 T cells. **i**, central memory CD8 T cells from spleens of Balb/c mice (n= 3-5) bearing K7M2 pulmonary sarcomas harvested 24h after a third weekly dose of i.v. RNA-LPA loaded with tumor derived mRNA (from K7M2). **j**, C57Bl/6 mice (n=7-8) were implanted with B16F10-OVA subcutaneously on day 0 and injected with OT-1 cells intravenously next day. Tetramer+ T cells from spleens were harvested 24h after third weekly dose of i.v. OVA specific RNA-LPA. **k**, Survival curve of Balb/c mice (n= 8) bearing pulmonary K7M2 sarcomas treated with i.v. tumor derived RNA-LPA. **l**, Survival curve of Balb/c mice (n= 5-8) bearing pulmonary K7M2 sarcomas treated with three weekly i.v. GFP or K7M2 tumor derived RNA-LPA (data is aggregate of two separate experiments). **m**, Re-challenge of long-term surviving Balb/c mice (n=7-8) previously treated with K7M2 tumor derived RNA-LPA inoculated at 120 days with K7M2 i.v. versus a new cohort of untreated mice. **n**, Survival curve of SCID Fox-Chase mice on Balb/c background (n= 7-8) bearing pulmonary K7M2 sarcomas treated with three weekly i.v. GFP or K7M2 tumor derived RNA-LPA. **o**, Survival curve of C57Bl/6 mice (n=14) bearing intracranial KR158b gliomas transduced with luciferase and pp65 and treated i.v. with weekly GFP or pp65 RNA-LPAs. **p**, Survival curve of C57Bl/6 mice (n= 7-8) implanted neonatally (P1) with K2 midline gliomas expressing H3K27M and treated with H3K27M encoding RNA-LPA. Significance was determined via parametric student’s t-test (**f-j**), and log-rank test (**k-p**). Error bars are reported as the standard error of the mean (**f**) and standard deviation of the mean (**g-j**).

We tested different RNA-LPA ratios for kinetics of aggregate formation and immunogenicity. Increased particle concentration at fixed mRNA doses resulted in smaller aggregates (**Fig. 1e, Extended Data Fig. 1c**), which appear more lamellar and condensed by electron microscopy (**Extended Data Fig. 1d)**; surprisingly, these heightened interferon (IFN) responses after systemic (intravenous) delivery (**Fig. 1f**) and increased DC activation and central memory T cells in treated animals bearing K7M2 pulmonary sarcomas (**Fig. 1g-i**). Increasing LP:RNA ratio also enhanced antigen specific immunity against model antigens (**Fig. 1j**) and significantly lengthened long-term survivorship (**Fig. 1k)** in animals with metastatic pulmonary nodules. Although immunologic responses were superior at increasing LP:RNA ratios, survivorship was comparable between RNA-LPAs at 1:7.5 and 1:15 mass ratios suggesting these to be ideal for RNA-LPA formation. Therefore, we did not test additional LP-RNA ratios and prioritized testing and development of RNA-LPAs at 1:15 mass ratios. In follow-up experiments, we showed that the survival benefit from RNA-LPAs was dependent on total tumor-derived mRNA as opposed to a nonspecific mRNA control (GFP, **Fig 1l**). To demonstrate that RNA-LPAs induce antigen specific recall, we re-challenged long-term survivors from animals treated with K7M2 specific RNA-LPAs with tumor cells (K7M2 i.v.) and demonstrated that vaccinated animals could successfully ward off tumors (**Fig. 1m)**. Antigen specific recall responses were confirmed in ex vivo restimulation assays (**Extended Data Fig. 1e**). Finally, long-term survival from tumor specific RNA-LPAs was abrogated in SCID mice establishing requirement of antigen specificity and adaptive immunity for improved outcomes (**Fig. 1n**).

RNA-LPAs can be made from unfiltered total tumor derived mRNA (experiments above) or by encoding for synthetic overexpressed antigens or neoantigen epitopes. We tested whether RNA-LPAs encoding for antigens relevant in human cancers could elicit anti-tumor efficacy in murine models expressing these epitopes. RNA-LPA induced anti-tumor efficacy against the physiologically relevant antigen pp65 (overexpressed in glioblastoma) in animals bearing KR158b-luc-pp65 cortical gliomas (**Fig. 1o**). In a murine model for diffuse midline glioma (DMG) beginning treatment near endpoint, RNA-LPAs encoding for H3K27M (conserved mutation expressed in human DMG) also elicited long-term survivor benefits (**Fig. 1p)**. These results have implications for diffuse intrinsic pontine glioma (DIPG) which remains unresectable and uniformly fatal in children. Thus, in early and late disease models, RNA-LPA encoding for total tumor or tumor specific/associated antigens improves median and long-term survivor outcomes.

We next sought to determine mechanism of RNA-LPA activity and hypothesized that 200-500 nm aggregates would be entrapped in reticuloendothelial organs^10^. It has recently been shown that the spleen has an open microcirculation with intervening connective tissue between endothelial cells^11^ making it ideally suited to take up larger particle aggregates. We utilized tdtomato reporter mice under Cre-lox to visualize cellular localization of RNA-LPAs in spleen. Unlike prototypical mRNA therapies engineered to deliver antigen to macrophages or dendritic cells, Cre encoding RNA-LPAs localized to the interface of the splenic white pulp (**Fig. 2a, Extended Data Fig. 2a**) and elicited tdtomato expression in perivascular stromal cells (**Extended Data Fig. 2b**). Co-localization analyses reveal tdtomato expressing cells that are laminin positive, indicating they are fibroblastic reticular cells (FRCs) (**Fig. 2b-c**). We demonstrate that suspected RNA-LPA transfected FRCs are proximal to and have direct contact with F4/80+ APCs (**Fig. 2d-e**) and CD11b+ myeloid cells (**Extended Data Fig. 2c-d**). FRCs make up conduit systems in lymphoreticular organs to entrap pathogens and shuttle their delivery to APCs for induction of adaptive immunity.^12,13^ Under stress, human fibroblasts are known to secrete copious amounts of type I IFNs^14^, and after transfection with RNA-LPA (**Extended Data Fig. 2e, f**) induced chemokine response signatures for myeloid and lymphoid cell recruitment (**Extended Data Fig. 2g**). In vivo, within 6h of i.v. RNA-LPA, we observed rapid increases in serum cytokines including type I/II IFNs, IL-12, and TNF-α (**Fig. 2f**) and chemokines CCL2, CCL4, and CXCL9-10 (**Fig. 2g**) critical for recruitment of monocytes^15^ and lymphocytes.^16,17^ In concert with this response, RNA-LPAs elicited substantial recruitment of peripheral blood mononuclear cells (PBMCs) evidenced by transient monocytopenia and lymphopenia (**Fig. 2h, Extended Data Fig. 2h)**, which correlated with increases in activated DCs and T cells in lymphoreticular organs (**Fig. 2i, Extended Data Fig. 2i**). Surprisingly, these remodeling effects were not unique to the periphery, but could also reprogram the TME. In mice bearing malignant gliomas, we show that the TME undergoes significant inflammatory changes within 24h of tumor specific RNA-LPA with increased gene signatures for antigen presentation, IFN signaling, and cytotoxicity (**Fig. 2j, Extended Data Fig. 3a-e)**. Similar to the danger response and immune activation in the periphery, RNA-LPA increased gene signatures for damage associated molecular patterns (i.e. RPS13), pathogen recognition receptors (PRRs), antigen presentation, and myeloid/lymphoid activation in the TME **(Fig. 2j, Extended Data Fig. 3a-e**). Anti-tumor efficacy from RNA-LPA (**Fig. 2k)** and trafficking of lymphocytes to lymphoreticular organs (**Extended Data Fig. 3f**) were dependent on type I IFN; however, unlike other single stranded mRNA formulations^18^, RNA-LPA responses were not strictly dependent on TLR7 (**Fig. 2l**) or downstream MYD88 adapter protein (**Fig. 2m**). In contrast to TLR7, stimulation of RIG-I was critical for RNA-LPA mediated activity as responses were significantly attenuated in RIG-I knock-out mice (**Fig. 2n)**, suggesting that RNA-LPAs develop secondary structures that activate intracellular sensors for double stranded elements. Thus, unlike other mRNA vaccine formulations, RNA-LPAs function as immunomodulating agents that activate intracellular PRRs eliciting rapid mobilization of PBMCs that simultaneously reprogram both systemic immune cells and those within the TME.

**Fig. 2:**
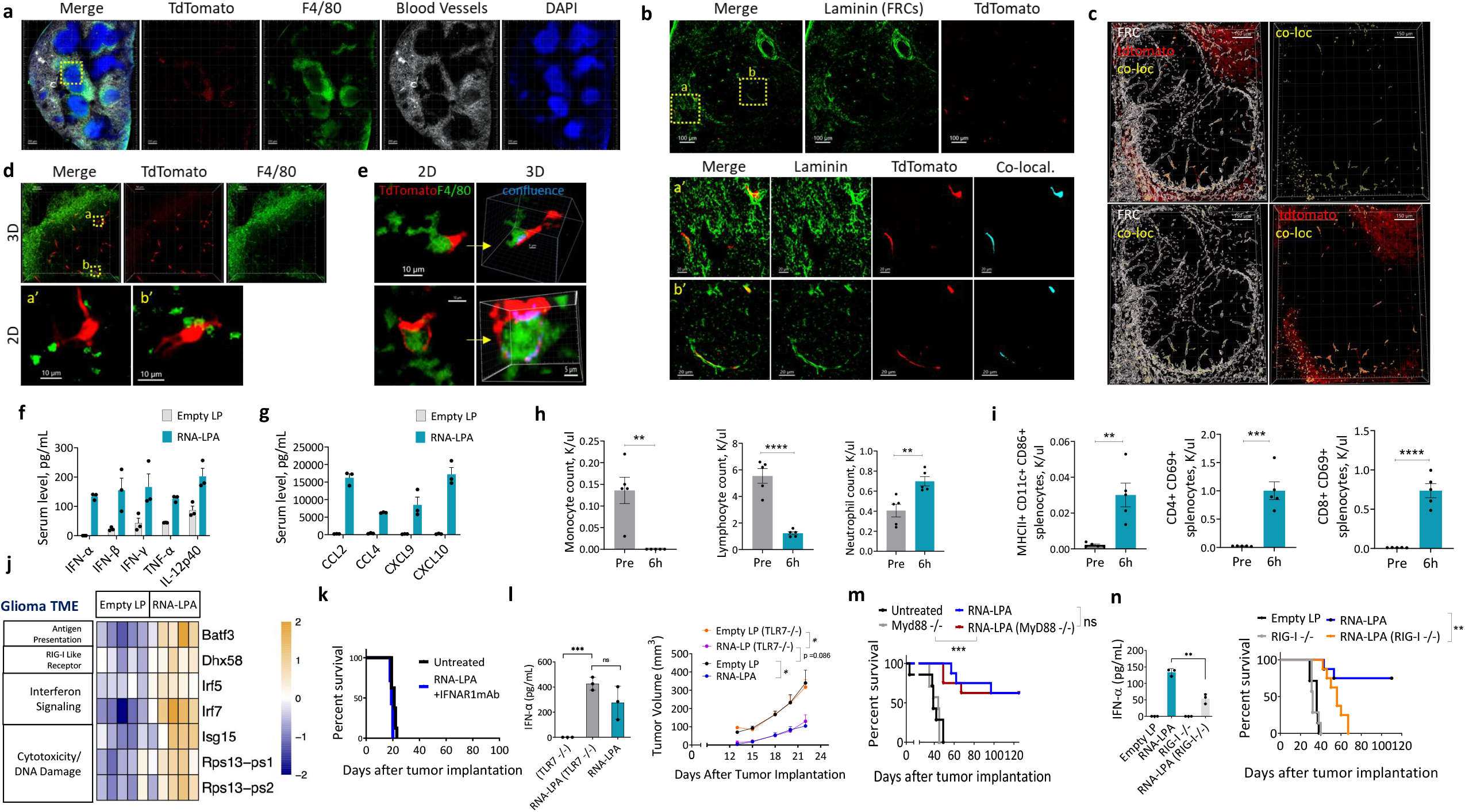
RNA-LPAs transfect lymphoreticular stroma, chemoattract PBMCs, reprogram the TME and mediate activity through stimulation of intracellular PRRs. **a**, Cross section of the whole spleen 2 days following i.v. injection with Cre RNA-LPA. Yellow dashed box maps to magnified panels ‘d.’ **b**, *Top row*, lower magnification of laminin (FRC marker) and tdtomato signal; *bottom rows* represent higher magnification of yellow boxed areas. Co-localization between laminin and tdtomato, calculated using voxel-based signal overlap, is displayed as an independent channel in light blue. **c**. 3D surface-based co-localization between FRCs (laminin+) with tdtomato. Colocalization is displayed as an independent channel in yellow. **d**, *Top row* maps to yellow box in panel ‘a’ with 3D overlay of tdtomato with F4/80; *bottom row* displays two inlaid boxes at higher magnification in 2D. **e**, *Left column* shows 2 different tdtomato cells (in 2D) interfacing with macrophages (separate from ‘d’); *right column*, 3D capture of these cells, rotating to display maximum contact ‘confluence’ points identified via voxel co-localization (blue) **f, g**, Cytokine/chemokine response panel from C57Bl/6 mice (n=3) treated with i.v. RNA-LPA. **h**, Absolute counts of peripheral white blood cells from C57Bl/6 mice (n=5) 6h after i.v. RNA-LPA. **i**, Absolute counts of activated DCs and activated T cells in spleens of C57Bl/6 mice (n=5) harvested 6h after i.v. RNA-LPA. **j**, RNA sequencing of established KR158b-luc intracranial tumors harvested 24h after a single tumor derived RNA-LPA. **k**, Survival curve of C57Bl/6 mice (n=7-8) bearing pulmonary K7M2 sarcomas treated i.v. with three weekly RNA-LPAs concomitantly with IFNAR1 mAbs. **l**, Serum analysis of IFN-α (left), n=3, and survival curve (right) from C57Bl/6 wild-type versus TLR7 knock-out mice (n=5-8) bearing subcutaneous B16F10-OVA melanomas treated i.v. with OVA RNA-LPA. **m**, Survival curve from C57Bl/6 wild-type versus MYD88 knock-out mice (n= 7-8) bearing pulmonary B16F10-OVA melanomas treated i.v. with OVA RNA-LPA. **n**, Serum analysis of IFN-α (left), n=3, and survival curve (right) from C57Bl/6 wild-type versus RIG-I knock-out mice (n=7-8) bearing pulmonary B16F10-OVA melanomas treated i.v. with OVA RNA-LPA. Significance determined via parametric student’s t-test (**h, i, l** (left), **n** (left)), Mixed effects models/repeat ANOVA models (**l**, right) and log-rank test (**l** (right), **m, n** (right)). Error bars reported as the standard error of the mean (**h, i, l** (right)) and the standard deviation of the mean (**l** (left), **n** (left)).

To translate this approach and validate its effects in a naturally occurring large animal model, we treated pet dogs with spontaneous gliomas, a terminal disease in canines with a median survivorship of less than 1-2 mo.^19-22^ We treated 10 canines, 5 subjects in Group A receiving RNA-LPAs bi-weekly following tumor biopsy, and 5 subjects in Group B first receiving neoadjuvant RNA-LPA (encoding an irrelevant antigen not expressed in canine glioma, pp65) before tumor biopsy and preparation of tumor specific RNA-LPA as in Group A (**Fig. 3a**). Tumor specific formulations were prepared from individualized biopsies following RNA extraction and mRNA amplification. In all canines, we observed analogous effects to mice, including rapid induction of pro-inflammatory cytokines (i.e. IFN-α, **Fig. 3b**) and chemokines (i.e. CCL2, **Fig. 3c**), and mobilization of PBMCs (**Fig. 3d-e**) within hours of first RNA-LPA administration. While monocytes nadir at 2h (**Fig. 3d, Extended Data Fig. 4a**), lymphocytes appear to a nadir at 6h post-infusion (**Fig. 3e, Extended Data Fig. 4b**); these results correlate with increasing temperatures in treated canines (**Fig 3f**). To determine whether RNA-LPAs could reprogram the TME of canine gliomas, we treated a cohort of canines (Group B) with neoadjuvant RNA-LPA prior to biopsy. Within days of an initial RNA-LPA, we observed rapid reprogramming of the canine glioma TME with increased gene signatures for antigen presentation, IFN signaling, and cytotoxicity (**Fig. 3g, Extended Data Fig. 4c**). Remarkably, we witnessed near complete resolution of tumor burden in one subject receiving adjuvant radiotherapy following neoadjuvant RNA-LPA (**Extended Data Fig. 4d**). This was the only reported subject receiving adjuvant therapy (post-biopsy) aside from supportive care and RNA-LPA. Compared to historical controls of subjects receiving supportive care alone (median survival 35 days^21^), RNA-LPA encoding for tumor specific mRNA improved median survival outcomes (median OS, 123) in subjects with spontaneous canine glioma (**Fig. 3h, Extended Fig. 4e**). While RNA-LPA administrations were generally well tolerated in canines, we corroborated safety in preclinical GLP toxicology studies. In naïve C57Bl/6 mice receiving RNA-LPAs encoding for multiple mRNA species (tumor amplified mRNA and pp65 mRNA RNA-LPA) there was no adverse gross or microscopic test article related findings at 35, 56 and 110 days (**Extended Data Fig. 5**).

**Fig. 3:**
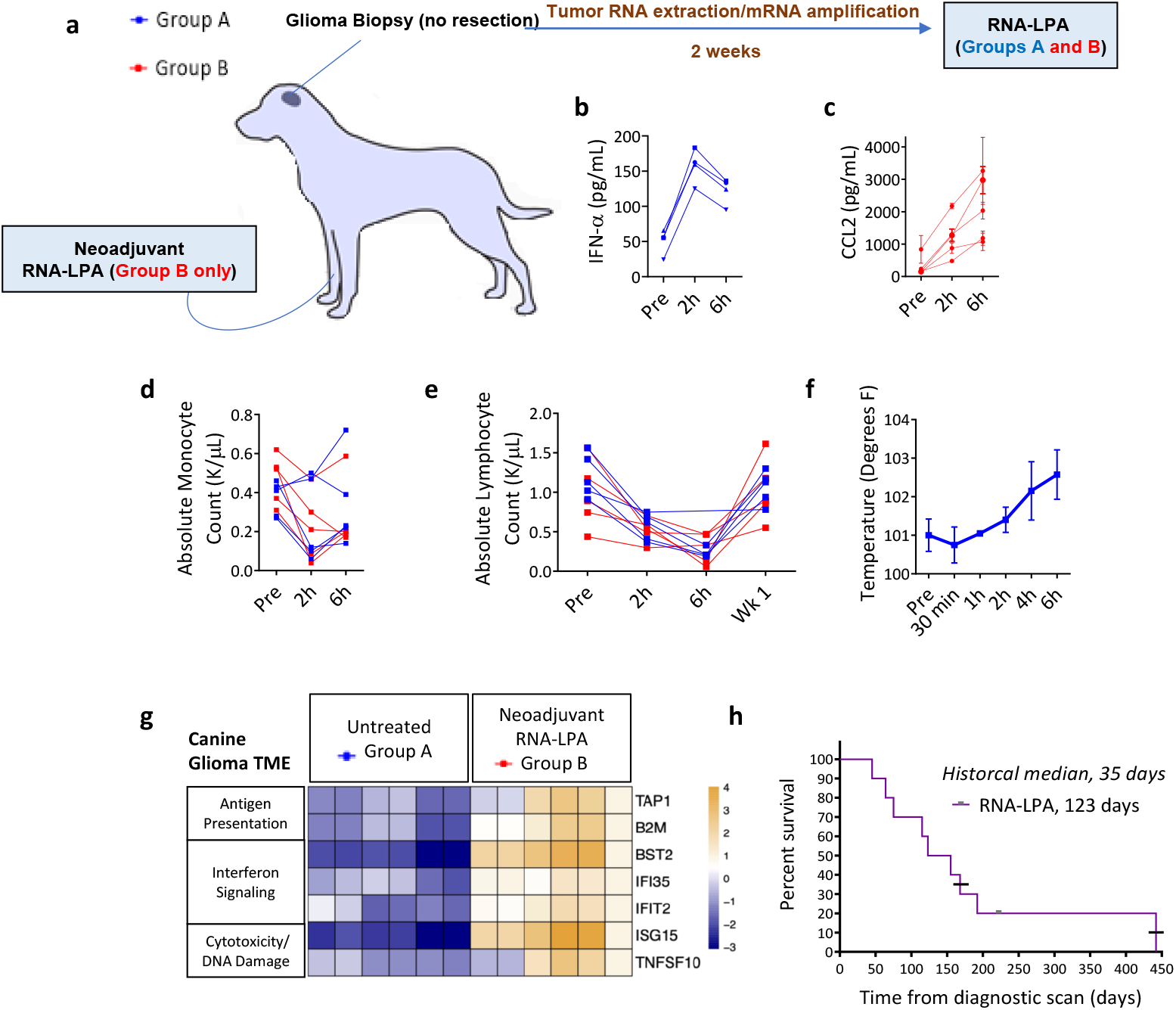
RNA-LPA mobilize PBMCs and reprogram the TME of client-owned canines with terminal gliomas. **a**, Study schema of client-owned canines with terminal gliomas enrolled to receive RNA-LPA. Group A: Adjuvant tumor derived RNA-LPA alone” vs. “Group B: Neoadjuvant pp65-RNA-LPA followed by tumor derived RNA-LPA. **b, c**, Serum from canines was obtained for analysis of IFN-α (Group A, n=4) (**b**) and CCL2 (Group B, n=5) (**c**). **d, e**, Absolute monocyte (**d**) and absolute lymphocyte (**e**) count from canines (Group A and B, n= 10) pre, 2, and 6h post-initial RNA-LPA. **f**, Temperature measurements from first 4 Group A canines following RNA-LPA (Subject 1-2, after initial infusion; Subject 3-4, after 3^rd^ infusion). **g**, Nanostring analysis of untreated canine glioma specimens (Group A, n=3) or within 48h of RNA-LPA (Group B, n=3). **h**, Survival curve of canines (Group A and B, n= 10) bearing intracranial gliomas undergoing treatment with RNA-LPAs. Crosses at 168 days and 442 days delineate 2 canines receiving additional therapies: pre-vaccine CCNU (168 days) and post-vaccine radiation therapy (442 days). Historical control for canines receiving only supportive care palliation (mean, 63 days; SD = 14 days).^21^ Error bars reported as standard error of the mean (**c, f**) from individual subject replicates (**c**) and pooled replicates across canines (**f**)

Under FDA-IND (BB-19304), we conducted a first-in-human (NCT04573140) phase 0 (acceleration titration design) dose study in 3 patients evaluating the feasibility, safety, and activity of RNA-LPA targeting pp65 mRNA (tumor associated antigen expressed in glioblastoma^23-27^) and autologous tumor mRNA (**Fig. 4a**) in patients with primary MGMT unmethylated glioblastoma. After surgery and chemoradiation, patients received up to 3 biweekly (every 2 weeks) doses of RNA-LPA followed by a monthly booster. Patients were treated with mRNA infusion doses of 0.625-1.5 μg/kg. In all patients, we observed significant and rapid increases in pro-inflammatory cytokines (**Fig. 4b-c, Extended Fig. 6a**) and chemokines (**Fig. 4d, Extended Fig. 6a**). All patients developed immune-related symptoms 2-6h post-infusion (e.g. low-grade fever, nausea, chills, rigors) that defervesced within 24-48h. Cytokine release correlated with mobilization of PBMCs beginning with monocytes at hour 2 (**Fig. 4e**), followed by lymphocyte nadir and neutrophilia at hour 6 (**Fig. 4f-g**). Using multi-parameter flow cytometry (**Extended Data Fig. 6-7**), we demonstrate that antigen presenting cells (HLA-DR expressing) rapidly decrease from peripheral blood (**Fig. 4h**) suggesting recruitment to lymphoreticular organs. While CD11c cells rapidly decrease (**Fig. 4i**), there is an increase in: 1) percent circulating plasmacytoid DCs (**Fig, 4j**); 2) early activation of CD8 lymphocytes (**Fig. 4k**); and 3) expansion of antigen specific T cell responses (**Fig. 4l-o, Extended Data Fig. 7b**). Using sequencing of T cell receptors pre- and post-infusion, we show that TCR repertoire of the most prevalent clonotypes changes by ∼10% in Patients B42 and E35 after 1-2 infusions and nearly 20% in Patient A25 after 4 infusions (**Extended Data Fig. 8a-c**) highlighting the ability of RNA-LPAs to reprogram adaptive immune repertoires. Patients A25 and E35 had increased progression free survivorship compared with historical median of 6 mo^28^ and patient B42 has begun long-term follow-up.

**Fig. 4:**
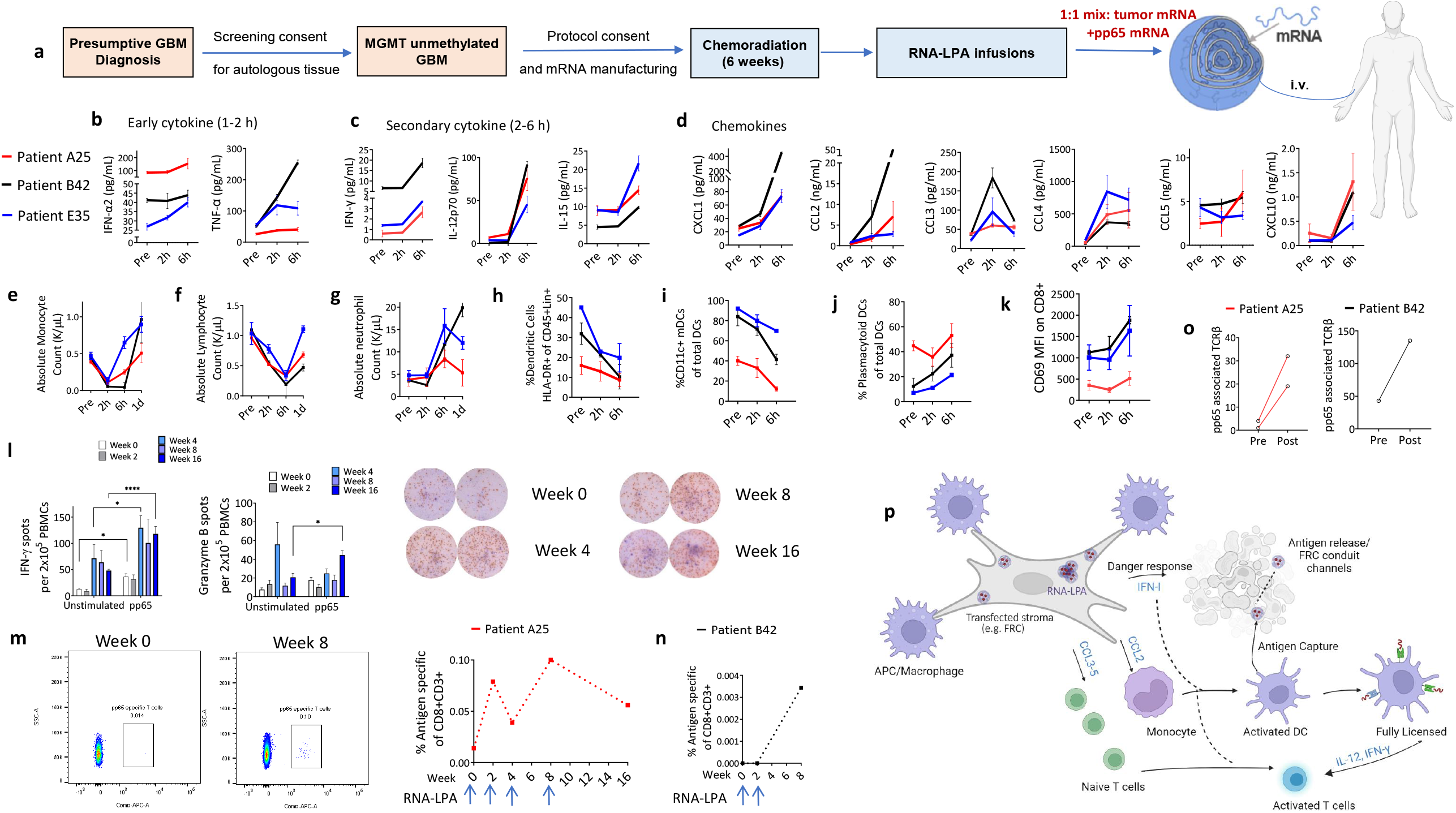
RNA-LPA induce rapid cytokine/chemokine release, PBMC mobilization, and antigen specific T cell responses in human glioblastoma patients. **a**, Study schema of MGMT unmethylated glioblastoma patients enrolled to receive RNA-LPA. **b, c, d** Cytokine/chemokine sera measurements pre, 2, and 6h post-RNA-LPA. **e, f, g**, Absolute counts of peripheral blood monocytes (**e**) lymphocytes (**f**), and neutrophils (**g**). **h, i, j**, Fraction of APCs (**h**) and DC subtypes including mDCs (**i**) and pDCs (**j**). **k**, Mean fluorescent intensity (MFI) of activated CD8 T cells. **l**, IFN-γ and granzyme B Elispots using unstimulated and pp65 re-stimulated PBMCs from Patient A25. **m, n**, Flow cytometric analysis of antigen specific T cells in Patients A25 and B42 by tetramer staining for HLA-A2 restricted pp65 epitope. **o**, Significantly expanded pp65-specific TCRβ in Patient A25 and B42 samples after 2 and 4 RNA-LPA infusions respectively. **p**, Proposed mechanisms for RNA-LPA mediated innate and adaptive mobilization/activation (Image created with Biorender.com).

Messenger RNA vaccines have been developed for prevention of infectious diseases, providing a blueprint for the development of therapeutic cancer vaccines. These blueprints are predicated on LNP designs that are relatively inert at physiologic pH (with cholesterol, PEG, and helper lipids) to promote gene expression of target antigens like SARS-CoV-2 full-length spike protein.^29^ Similarly, mRNA constructs are modified with pseudouridines to silence innate stimulation and maximize gene expression and adaptive immunity. However, unlike spike protein in the prophylactic setting, cancer antigens are already expressed and typically mediate tolerogenic responses.^30^ In these settings, adaptive immunity is stymied by a hostile TME leading to poor T cell trafficking, penetration, and persistence. These challenges are salient for cancer vaccines, checkpoint inhibitors, and CAR T cells. Overcoming them requires new technologies that make cancer antigens look more dangerous while simultaneously reprogramming the TME.

The current paradigm of mRNA delivery is via payload packaging into the core of LNPs for endocytic delivery, TLR stimulation and unpackaging. However, natural designs have revealed an exquisitely simple and highly effective approach to making payloads appear more dangerous.^16,31-38^ Recent work with high-resolution imaging has revealed that viruses packaged together into vesicles are significantly more infectious than a single virus.^16,31-38^ Mimicking this structure, we have developed a dangerous appearing vaccine that utilizes tumor mRNA wrapped into a multi-lamellar vesicle within an onion-like hierarchical structure.

By using mRNA as a molecular bridge with cationic liposomes, self-assembling lamellar aggregates form that mimics infectious emboli upon intravenous administration. Analogous to highly potent viral vesicles, RNA-LPAs elicit rapid augmentation of innate and adaptive immune responses following a danger response that recruits and activates the bulk of circulating PBMCs to sites of particle localization (**Fig. 4p**). While single-stranded mRNA vaccines rely on TLR engagement, RNA-LPAs activate intracellular PRRs such as RIG-I. By aggregating, mRNA secondary structures are likely enabled that stimulate double-stranded sensing PRR machinery. Critically, this activation rapidly resets the innate immune system in both the periphery and TME unlocking therapeutic T cell activity. Our approach can incorporate synthetic tumor-associated or patient-specific total tumor-derived mRNA into multilamellar LPAs enabling the immune system to parse the entire cancer transcriptome.

In our first treated human subjects, we observe massive recruitment of PBMCs, rapid activation of DCs /T cells and expansion of antigen specific T cell responses. These data support the use of RNA-LPAs as novel tools to restore response in poorly immunogenic tumors and challenge the paradigm that mRNA cancer vaccines must be engineered as nanoparticles to target DCs. Alternatively, mRNA aggregates mimicking dangerous emboli localize to lymphoreticular organs activating intracellular PRRs for the recruitment of immune cells that mediate rapid and enduring cancer immunotherapy.

## Data Availability

All data produced in the present study are available upon reasonable request to the authors

## Figure Legends

**Extended Data Fig. 1: RNA-LPA characterization. a**, Size distribution of RNA-LPAs (n=5) by Nanosight imaging. **b**, Zeta potential measurements of RNA-LPAs at fixed mRNA doses with increasing LP ratio. **c**, Size distribution (as measured by the radius of gyration, Rg) 15 min post-mixing. **d**, Cryo-electron micrographs of Empty LPs or different ratios of mRNA lamellar aggregates. **e**, Memory recall response from Balb/c mice (n= 4-5) splenocytes co-cultured with K7M2 versus control tumor (B16F0) after three weekly i.v. K7M2 tumor derived RNA-LPA. Significance determined via mixed-effects analysis and error bars reported as standard deviation of the mean (**e**).

**Extended Data Fig. 2: RNA-LPA transfection in vitro and in vivo. a**, Microscopy of spleens from cre-lox tdTomato reporter mice treated with i.v. Cre RNA-LPA. **b**, 3D and magnified 2D image of vasculature in splenic white pulp demonstrating that tdtomato+ cells are perivascular. **c**, tdtomato+ FRCs in close contact with CD11b+ cells. **d**, mp4 clipping plane fly-through of panel c. **e**, Light and fluorescent microscopy of RNA-LPA transfected human FRCs in vitro. **f**, Flow cytometric analysis of RNA-LPA transfected human FRCs. **g**, Serum ELISA from RNA-LPA transfected FRCs. **h**, Absolute blood counts over time in mice treated with RNA-LPA (n=5). **i**, Absolute counts over time for DCs and activated T cells in spleens of mice treated with RNA-LPA. Error bars reported as standard error of the mean (**f, g, h, i**).

**Extended Data Fig. 3: RNA-LPAs reprogram the TME and elicit IFNAR1 dependent lymphocyte trafficking. a, b, c, d, e**, RNA sequencing of established KR158b-luc intracranial tumors harvested 24h after a single tumor derived RNA-LPA, compared to Empty LP treated tumors. Heatmaps of differential gene expression showing fold change in gene expression, with yellow indicating upregulation and blue indicating downregulation: DNA damage response (a, p-Adj value = 0.02074), Pathogen Recognition Receptor Signaling (b, p-Adj = 0.05285), Lymphocyte Activation (c, p-Adj = 0.02760), Myeloid Leukocyte Activation (d, p-Adj = 0.00761), Antigen Processing and Presentation of Peptide Antigen (e, p-Adj = 0.04409). (**f**) Absolute monocyte and lymphocyte count in animals within 24h of RNA-LPA treatment in wild-type and IFNAR1 knock-out animals. Significance determined via parametric student’s t-test and error bars reported as standard error of the mean.

**Extended Data Fig. 4: RNA-LPA responses in client-owned canines**. Absolute monocyte (**a**) and lymphocyte (**b**) count averaged from canines (n= 10) pre, 2, and 6h post-RNA-LPA. **c**, Heatmaps displaying differential gene expression of Antigen Presentation, Cytotoxicity and Interferon Signaling pathways using nCounter Canine IO Panel from Nanostring. Canine Glioma tissue specimens were collected at 48h after RNA-LPA treatment (n=3) compared to untreated canine glioma specimens (n=3). Yellow indicating upregulated genes, and blue indicating downregulated genes, with p-Adj value ≤0.05. **d**, MRI images of canine glioma receiving neoadjuvant RNA-LPA before (left) and after adjuvant radiotherapy. **e**, Kaplan-Meier curve of canines (Group A and B) receiving RNA-LPA compared with historical control of canines receiving supportive care.

**Extended Data Fig. 5: RNA-LPA toxicology studies. a**, GLP toxicology study groups. Treatment groups consisted of Empty LPs or LPAs of tumor mRNA (amplified from KR158b) and pp65 mRNA mixed 1:1. **b**, Study schema of GLP toxicology report summary. **c**, representative H&E analysis of organs from non-clinical toxicology studies.

**Extended Data Fig. 6: RNA-LPA cytokine response panel in human glioblastoma patients and gating strategy for analysis of DC subsets. a**, Cytokine/chemokine sera measurements pre, 2, and 6h post-RNA-LPA. **b**, Gating strategy for analysis of DC subsets.

**Extended Data Fig. 7: Gating strategy for analysis of lymphocyte subsets and CMV associated TCRβ expansion. a**, Human lymphocyte gating strategy. **b**, Number of productive complementarity-determining region 3 (CDR3) templates detected in clinical trial patients pre- and post-RNA-LPAs (Post samples collected after 4 infusions in Patient A25, after 2 infusions in Patient B42, and after 1 infusion in Patient E35).

**Extended Data Fig. 8: TCRβ spectratyping. a, b, c**, Differential abundance of productive TCRβ templates pre- and post-RNA-LPA in the first three patients. Orange circles represent TCRβ counts that are significantly higher than pre-infusion TCRβ templates. (Post samples collected after 4 infusions in Patient A25, after 2 infusions in Patient B42, and after 1 infusion in Patient E35).

## Acknowledgements

The authors would like to thank Gabriel De Leon for assistance with generating KPGL cell line; Lana Fagman and Kaylee Young for veterinary trials assistance; Tina Caton, Kathryn McKibben, Alexa Krasnogorska, Ievgenii Krasnogorskyi for product manufacturing assistance; Alicia Wyrick and Renee Boyette for administrative financial assistance; Kelly Hitchner for human clinical trial coordination; Jenna Weight, Cassie Kline, Tab Cooney and PNOC consortium for human trial oversight and data/safety monitoring.

## Funding

This work was supported by federal awards W81XWH-17-1-0510, K08CA199224, R37CA251978, R01CA266857, R01FD007268 (FDA – OOPD, Office of Orphan Products Development); Florida Department of Health 20B11 (Bankhead Coley) and 20L07 (Live Like Bella) awards; and foundation grants from CureSearch (Catapult Award), The Rally Foundation, Hyundai Hope on Wheels (Hope-Scholar Award), St. Baldrick’s, Chance for Life, Team Jack, Stop Children’s Cancer, Danny’s Dream, and The National Pediatric Cancer Foundation. Funding is also supported in part by the University of Florida Interdisciplinary Center for Biotechnology Research with funding provided by NIH Grant (1S10OD020026). J.G. acknowledges support R35GM146877 from NIGMS.

## Conflicts of Interest

D.A.M. holds ownership interest in iOncologi, Inc. The manuscript discusses patented technologies from H.M.G, P.C., S.Q., J.M., A.G., J.H., W.G.S., M.R., D.A.M., and E.J.S. Patented technologies are under option to license by iOncologi, Inc. The other authors declare no conflicts.

## Materials and Methods

### mRNA

Synthetic mRNAs encoding for full-length model antigens (EGFP, luciferase, and OVA albumin) were either purchased from Trilink Biotechnologies (Cat. L-7601, L-7602, and L-7610, respectively) or manufactured in-house using our custom DNA plasmids. Tumor associated antigen pp65-lysosome-associated membrane glycoprotein (LAMP)^23^ and histone K27M sequences were cloned in our custom plasmids. mRNA was produced from plasmid through a series of steps: First, the plasmid was transformed into bacterial cells and grown in culture. A maxi-preparation was performed to isolate the plasmid, which was then digested with restriction enzymes to linearize it. The linear plasmid was purified and used as a template for in vitro transcription (IVT) using a commercial kit (ThermoFisher Scientific, cat. AM1344). The resulting mRNA was subsequently purified using RNA purification kits (Qiagen, cat. 75162 and 74104). Total tumor derived mRNA was manufactured from total RNA extracted from parental cell lines (K7M2, K-luc, B16F0) or primary tumor samples (canine/human) using Qiagen kits. RT-PCR was used to make a cDNA library (Takara, cat. 639537 and 639202) before the use in IVT reactions and RNA clean-up for purified mRNA antigens. The quality of the mRNA was assessed as follows: Concentration and purity of the mRNA were determined using a Nanodrop spectrophotometer; gel-electrophoresis and bioanalyzer analyses were performed to confirm the integrity and size distribution of the mRNA.

### RNA-LPA generation

Raw material for lyophilized DOTAP (1,2-dioleoyl-3-trimethylammonium-propane (chloride salt) was obtained from Avanti Polar Lipids (cat. 890890P). To develop pure liposomes and prevent ambient oxidation, we utilized a rota-evaporator system to remove solvents and rapidly reconstitute lipids in an aqueous solution. Briefly, chloroform was added to lyophilized DOTAP starting material and carefully disrupted through gentle rotation and sterile vacuum suction before lipid-film was re-suspended in PBS for rotational water bath heating, sonication and extrusion using 0.45 μm and 0.2 μm filters. After positively charged bilamellar LPs were formed, we added negatively charged synthetic mRNAs or tumor amplified mRNA as a molecular bridge to aggregate LPs at different mass ratios of RNA:LP, including 1:1, 1:3.75, 1:7.5 and 1:15 ratios. RNA-LPAs were validated based on concentration/size by Nanosight NS300 (Malvern Panalytical), zeta potential by Zetasizer Ultra (Malvern Panalytical), and encapsulation efficiency by gel-electrophoresis.

### Cryo-electron microscopy of LPs and RNA-LPAs

Cryo-electron micrographs were obtained by using core instruments via the Interdisciplinary Center for Biotechnology Research at UF. Samples of LPs and various ratios of RNA-LPAs were processed as previously described.^9,10^

### Fluorescent Imaging of RNA-LPAs

Prior to making mRNA-LPAs for imaging, LPs were labeled with CM-DiI Dye (Thermo Fisher Scientific, cat. C7000) at a final dye concentration of 1.8 μM. To fluorescently label mRNA, PBS was mixed with Trilink EGFP mRNA in equal volume ratios. To this, a 25x dilution (in PBS) of Green Fluorescent Nucleic Acid Stain (SYTO) (ThermoFisher Scientific, cat. S34854) was added and thoroughly mixed. Non-specific labeling by SYTO was ruled out by adding SYTO to both labeled and unlabeled LPs and imaging in the GFP and Cy3 channels. To ensure there was negligible channel bleed-through, imaging was done in the GFP and Cy3 channels on unlabeled LPs with SYTO-labeled mRNA and CM-Dil-labeled LPs with unlabeled mRNA. To prepare RNA-LPAs for imaging, labeled LPs were added to an 8-well plate (Cellvis) and image acquisition conditions were then set (see Image Acquisition Conditions). Following this, the labeled mRNA mixture was added, and imaging was started immediately.

#### Image Acquisition Conditions

Imaging was performed using a Nikon Eclipse Ti2-E and X-Cite XLED1 Multi-Triggering LED Illumination System. Automated image acquisition was controlled by Micro-Manager.^39^ Imaging was performed in the GFP and Cy3 channels (both at 20% full intensity at exposure times of 20 ms) over a period of 30 minutes in 1-minute intervals, with 3×3 tiling (tiles separated by 300 μm) and z-stacks at 0 μm (bottom of well), 25 μm, 50 μm, 75 μm, and 100 μm.

#### Image Segmentation and Analysis

Image segmentation with custom scripts was carried out entirely in the open-source image analysis software, FIJI (1.5.3q).^40^ Cy3 images were first preprocessed using a morphological top-hat filter and then a Gaussian filter to reduce background and noise. Automatic thresholding and segmentation were then performed. Identified clusters were then overlaid with the original images to take measurements. All resulting measurement files were then exported (as .csv files) for further analysis.

### Mice

All mouse experiments and procedures were approved by the University of Florida’s Institutional Animal Care and Use Committee (IACUC). Naïve mice including Balb/c (K7M2 studies) and C57Bl/6 mice (B16F0, B16F10-OVA, K2, KR158b-luc, KR158b-pp65-GFP-luc studies) were purchased from The Jackson Laboratory. Ai14 mice (or tdTomato cre-lox reporter mice, cat. 007914), TLR7 (cat. 008380), Myd88 (cat. 009088), RIG-I (cat. 046070-JAX) and INFAR (cat. 032045-JAX) knock-out mice were purchased from The Jackson Laboratory. OT1 transgenic (C57Bl/6-Tg(TcraTcrb)1100Mjb/J) mice were provided by UF ACS breeding facility. All mice were vaccinated intravenously with 25 μg/dose of RNA-LPA (mRNA fraction). Unless stated otherwise, all mice received RNA-LPAs at an RNA:LP mass ratio of 1:15. We administered anti-Interferon alpha receptor (IFNAR1mAb, Bio X Cell, cat. BE0241) antibody intraperitoneally (IP) at an initial dose of 500 μg/mouse once in the first week, followed by a maintenance dose of 250 μg/mouse twice a week for the remaining treatment period.

### Cell lines and implantations

K7M2 was obtained from ATCC (cat. CRL-2836) and inoculated via i.v. tail vein injection (K7M2: 1,250,000 cells) to form pulmonary tumors. B16F10-OVA (1 million cells, subcutaneous (s.c.) and 300,000 cells for i.v. implantations) and KR158b-luc (10,000 cells, intracranial (i.c.) implantation were obtained as kind gifts as previously described.^9,10^ KPGL was made through viral transduction of KR158b-luc tumor cells to express both pp65 and GFP. K2 tumor cells were obtained as a kind gift from Dr. Oren Becher at Northwestern University. In experiments with midline K2 tumors, neonatal mice were implanted midline and allowed to mature to mimic pediatric disease before vaccination. B16F0 was obtained from ATCC (cat. CRL-6322) and the human fibroblastic reticular cell line (FRC) was obtained from CancerTools.org (cat. 156505). All cell lines used in this study were cultured in accordance with the recommendations provided by the supplier.

### FRC transfection in vitro

Fibroblastic reticular cells (FRCs) were cultured in Fibroblast Medium containing 2% fetal bovine serum, 1% fibroblast growth supplement, and 1% antibiotic solution (SciencCell, cat. 2301). The cells were plated in a 6-well plate at a density of 150,000 cells per well for fluorescent microscopy analysis and 1,000,000 cells per well for flow cytometry analysis. Each well was treated with GFP RNA-LPA at a dose of 5 μg mRNA per well. Cells were maintained in an incubator at 37°C and 5% CO2 for 24 hours. The next day, cells were imaged and collected for flow cytometry analysis. Untreated cells were collected with Accutase (Innovative Cell Technologies, cat. NC9839010) for 3 minutes at 37°C and treated cells were collected with 0.25% trypsin-EDTA (Sigma-Aldrich, cat. T4049) for 5 minutes at 37°C. Flow cytometry was performed using the FACSymphony A3 (BD Biosciences) cell analyzer system and the FACSDiva data acquisition software (v9.1; BD Biosciences). The flow cytometry results were analyzed using FlowJo™ v10.8.2 Software (BD Life Sciences).

### Antigen Recall Assay

One week after the last RNA-LPA vaccine, splenocytes were collected for antigen recall assay. Mice were humanely euthanized in a CO2 chamber and spleens were collected using a standard aseptic technique. Single splenocyte suspension after passing homogenized spleen through 0.70um filters. Cells were centrifuged at 500g 5min at room temperature. The cell pellet was resuspended with 5ml of lysis buffer (BD, cat. 555899) for 3 min at 37°C. Lysis buffer was neutralized by two volumes of complete T cell medium and centrifuged at 500g for 5 min at room temperature. Complete T cell medium contains RPMI 1640 (Gibco, cat. 11-875-119), 10% fetal bovine serum (ThermoFisher Scientific, cat. 35-011-CV), 1% penicillin/streptomycin (Gibco, cat. 30-002-CI), 1% MEM non-essential amino acids (NEAA, Gibco, cat. 11140050), 1% sodium pyruvate (Gibco, cat. 11360070), 0.1% beta-mercaptoethanol (BME, Gibco, cat. 21985-023) without cytokines. Next, T cells and tumor cells were counted and plated at a ratio of 10:1 (effector:target) in a 96-well round bottom plate. Co-culture was maintained for 48 hours in an incubator at 37°C and 5% CO2.

### CBCs and cytokine analysis in mice

After collecting blood from mice, a portion was sent to the Clinical Pathology Laboratory at the University of Florida College of Veterinary Medicine for CBC (complete blood count) analysis with differential. Another portion was used to isolate serum through centrifugation at 10,000g for 10 minutes. Serum was then sent to Eve Technologies for multiplex cytokine analysis.

### Flow cytometry of mouse samples

Blood and spleens were collected from animals and digested as previously described to remove red blood cells and purify white blood cells for analysis of immune cell populations by flow-cytometry.^9,10^ The antibody panels employed were as follows: For Fig. 1g: CD45-PErCP-Cy5.5 (eBioscience, cat. 45-0451-82), CD11c-APC (eBioscience, cat. 17-0114-82), MHCII-FITC (eBioscience, 11-5321-85), CD86-BV421 (Biolegend, cat. 105032). Live/Dead Near-IR (Thermo Fisher Scientific, cat. L349750). For Fig. 1h and 1i: CD3-PE (BD, cat. 553064), CD4-BV510 (BD, cat. 563106), CD8-FITC (eBioscience, cat. 11-0081-85), CD44-PerCP-Cy5.5 (eBioscience, cat. 45-0441-82), CD62L-APC (BD, cat. 553152). Live/Dead Near-IR. For Fig. 1j: CD3-BV510 (BD, cat. 563024), CD8-FITC, OVA-tetramer-PE (MBL International, cat. TB-5001-1) and Live/Dead Near-IR. For Fig. 2i, left panel: CD45-PErCP-Cy5.5, CD11c-PE (Thermo Fisher Scientific, 2-0114-82), MHCII-FITC, CD86-BV510 (Biolegend, cat. 105039). For Fig. 2, middle and right panels: CD3-PE, CD4-BV510, CD8-FITC, CD69-APC (BD, cat. 560689). The flow cytometry results were analyzed using FlowJo™ v10.8.2 Software (BD Life Sciences).

### Immunofluorescence of mouse spleens

Microscopy of spleens in TdTomato reporter mice was performed as follow: tdTomato mice were injected with Cre RNA-LPA. 2 days later, mice were perfused with clarity solution. Spleens were processed by two different methodologies:

#### CLARITY 3D tissue clearing methodology

tissues were collected following cardiac perfusion with cold saline; this was subsequently followed by PBS supplemented with 4% acrylamide (Sigma-Aldrich, cat. A8887), 0.05% N,N’-methylenebisacrylamide (Sigma-Aldrich, cat. M7279), 4% paraformaldehyde, 0.25% VA-044 (TCI, cat. A3012), and 0.5 mg/mL Alexa Fluor 647 conjugated fixable Dextran 10,000 MW (Thermo Fisher Scientific, D22914). For 3D microscopy, tissues were stored at 4°C for three days, which allowed hydrogel permeation of tissues. Following hydrogel polymerization at 37°C x3 hours, tissues were passively cleared over three to seven days with PBS which contained 200 mM boric acid (Sigma Aldrich, cat. B0394) and 4% sodium-dodecyl-sulfate (Thermo Fisher Scientific, cat. BP166), pH 8.5 at 50°C. After clearing, samples were washed in PBS with 0.1% Triton X-100 for 2 days, blocked in PBS blocking buffer with 0.3% Triton X-100 (Thermo Fisher Scientific, cat. 85111), and immunolabeled at 4°C for 2 days with F4/80 (Cell Signaling, cat. 70076). Samples were washed in PBS with 0.1% Triton X-100 for 2 days followed by secondary immunolabeling with anti-Rabbit Alexa Fluor 488 (ThermoFisher Scientific, cat. A11034) plus DAPI (Sigma Aldrich, cat. MBD0015). Following a final wash in PBS with 0.1% Triton X-100 for 2 days, 62% 2,2’-Thiodiethanol (Sigma Aldrich, cat. 166782) was used for refractive index matching. Tissues were whole mounted onto slides for imaging using a Leica Stellaris 5 Confocal Microscope or Nikon A1RMP Confocal microscope. Image analysis was performed using Imaris x64 v9.7.0 software.

#### Formalin-sucrose-OCT methodology

tissues were collected and fixed in 10% formalin overnight at 4°C with agitation. The tissues were then cryoprotected by successive immersion in 15% and 30% sucrose solutions overnight at 4°C. Next, the organs were embedded in OCT solution (Sakura, cat. 4583) and frozen at -80°C. Tissue sections were cut in a cryostat at a thickness of 30-60 μm and mounted on slides. For immunostaining, sections were permeabilized and blocked with a solution containing 0.3% triton X-100, normal goat serum (NGS, ThermoFisher Scientific, cat. NC9660079), and phosphate-buffered saline (PBS) for 1 hour at room temperature. Anti-laminin primary antibody (Abcam, cat. Ab11575) was applied overnight at 4°C, followed by washing in PBS and incubation with anti-rabbit anti-Rabbit Alexa Fluor 488 for 2 hours at room temperature. Finally, the sections were mounted with mounting media (Vector Laboratories, cat. H-1700-10) and coverslipped. Imaging was performed using an Olympus IX70 for fluorescence images and the Nikon A1RMP Confocal microscope for confocal images.

### RNA sequencing (mouse and canine)

Fresh mouse tissue was collected and placed in RNAlater (Thermo Fisher Scientific, cat. AM7020) overnight at 4°C. The next day, RNAlater was removed and the tissue was cut into small pieces and frozen at -80°C until RNA extraction. RNA from the tissue samples was extracted using RNeasy Mini Kit (Qiagen, cat. 74104) and RNase-Free DNase Set (Qiagen, cat. 79254). RNA samples were then sent frozen to Novogene, who processed the samples as follows: mRNA enrichment was performed by Poly(A) capture or rRNA removal, followed by RNA fragmentation using Covaris or enzyme digestion. Then, cDNA reverse transcription and RNA library preparation were performed. The sequencing was carried out using the Illumina platform and the PE150 strategy. For RNA analysis, the software used was Mapping STAR v2.6.1 and Quantification HTseq v0.6.1. Differential analysis was performed using the Rosalind Platform (https://www.rosalind.bio/). Heatmaps were generated using R Studio.

Canine biopsies were collected and preserved in RNAlater (Thermo Fisher Scientific, cat. AM7020) overnight at 4°C to stabilize RNA and prevent degradation. The next day, RNAlater was removed and the tissue was cut into small pieces and frozen at -80°C until RNA extraction. Approximately 50mg of tissue was collected from each biopsy and processed for RNA extraction. Prior to preservation, samples were cleaned of necrotic tissue to ensure that the extracted RNA represented viable tumor tissue. The tissue was minced in a low-temperature controlled environment to minimize RNA degradation. RNA was then extracted using TRIzol Reagent (Invitrogen, cat. 15596026) following the manufacturer’s protocol. The amount of reagents used was adjusted according to the tissue size/weight following the manufacturer’s guidelines to optimize RNA yield and purity. The extracted RNA was cleaned and purified using the PureLink RNA Mini Kit (Invitrogen, cat. 12183018A). Concentration and quality of extracted RNA were measured using a Nanodrop One C (Thermo Fisher Scientific), and RNA concentration was normalized to a consistent starting quantity following NanoString’s recommended guidelines. The nCounter Canine IO Panel (Nanostring, cat. 115000465) was used to measure the expression of approximately 800 genes across various immune response and cancer pathways. The panel is designed specifically for use with canine samples, allowing for the study of the interaction between the immune system and cancer in dogs. Samples were labeled and hybridized following the manufacturer’s instructions to generate gene expression data. The data were normalized using the manufacturer’s recommended housekeeping genes in the nSolver Analysis Software, which uses the geometric mean of the housekeeping genes to normalize the data and reduce variability between lanes and hence samples.

Differential gene expression was performed using the Rosalind platform (https://www.rosalind.bio/), which allows visualization of gene expression data across various pathways. Heatmaps were then generated using R Studio (ggplot2; pheatmap packages) to visualize the differential gene expression patterns across the experimental groups.

### Canine trial

Client-owned canines diagnosed radiographically with glioma were enrolled in clinical trials through the owner’s consent. Subjects underwent glioma biopsy where total RNA was extracted, cDNA library generated and mRNA amplified following in vitro transcription reaction as described above. The first five dogs received 3 RNA-LPA (0.05 μg/kg/dose, mRNA fraction at 1:15 ratio with LPs) infusions one week apart starting two weeks after the tumor biopsy. The second five dogs (Group B) received a neoadjuvant non-specific RNA payload, pp65 mRNA, one day prior to biopsy, followed by three personalized mRNA vaccines one week apart starting two weeks after the biopsy. Blood samples were collected prior to infusion with RNA-LPA, at 2 and 6 hours post-infusion. A portion of the blood was used for complete blood count (CBC) analysis, while another portion was used for serum isolation and subsequent cytokine analysis. All analyses were conducted at the College of Veterinary Medicine at the University of Florida.

### Human trial

The phase 0 accelerated titration design (ATD) was embedded as a lead-in to an adult phase I trial for adult MGMT unmethylated glioblastoma patients. Potentially eligible participants were consented for eligibility screening through sterile collection of tumor that underwent RNA extraction, amplification, and loading of LPs. Data reported in manuscript is from first the three patients assigned random de-identified alpha numeric labels (A25, B42, E35). After surgical resection and confirmation of MGMT unmethylated status, patients were consented for trial enrollment. Radiotherapy began within 4 weeks (+/- 14 days) of surgery or sooner based on institutional preference. Forty-two doses of temozolomide 75mg/m2/day were given continuously during radiation for up to 49 days to account for delays in radiation treatment. RNA-LPA began within 4 weeks following radiation pending recovery of peripheral blood counts, and after assessment of post-radiation MRI (for baseline).

### Human Immunomonitoring

Blood was collected pre-RNA-LPA infusion, 2h post, and 6h post. Complete blood counts were performed through hospital labs, other measurements were obtained through separate blood collections. Serum was separated from peripheral blood cells and Multiplex Cytokine Assay analysis was done by Eve Technologies. Peripheral blood mononuclear cells (PBMCs) were isolated from RBCs by Ficoll density gradient separation before analysis by flow cytometry, Elispot, and TCR spectratyping.

#### Flow Cytometric Analysis

For human immunophenotyping by flow cytometry, two separate multicolor antibody panels were utilized. The first panel (Beckman Coulter, cat. B53309) was used for the identification of basic immune cell types and included the following markers provided in antibody panel kit, CD16-FITC (clone: 3G8), CD56-PE (clone: N901), CD19-ECD (clone: J3_119), CD14-PE-Cy7 (clone: RMO52), CD4-APC (clone: 13B8.2), CD8-Alexa Fluor 700 (clone: B9.11), CD3-APC-Alexa Fluor 750 (clone: UCHT-1), CD45-Krome Orange (clone: J33). Additionally, cells were stained with Live/Dead violet fixable viability stain (ThermoFisher Scientific, L34963), and the following antibodies from BD Biosciences PD-L1-BUV661 (clone: MiH1, BD, cat. 741666), HLA-DR-BV786 (clone: G46-6, BD, cat. 564041), and CD69-BUV737 (clone: FN50, BD, cat. 612817). The second panel (Beckman Coulter, cat. B53351) was used for the identification of dendritic cell subtypes and included the following markers provided in the antibody panel kit, CD16-FITC (clone: 3G8), CD1c-PE-Cy5.5 (clone: L161), CD11c-PE-Cy7 (clone: BU15), Clec 9A-APC (clone: 8F9), CD123-APC-Alexa Fluor 700 (clone: SSDCLY107D2), HLA-DR-Pacific Blue (clone: IMMU-357), CD45-Krome Orange (clone: J33), and a lineage gate with the following PE-conjugated antibodies CD3 (clone: UCHT-1), CD14 (clone: RMO52), CD19 (clone: J3-119), CD20 (clone: HRC20), CD56 (clone: N901). Additionally, cells were stained with Live/Dead violet fixable near-IR viability stain (Thermo Fisher Scientific, L34975), and the following antibodies from BD Biosciences PD-L1-BUV661 (clone: MiH1), CD80-BV786 (clone: L307.4, BD, cat. 564159), and CD86-BV786 (clone: FUN-1, BD cat. 740990). Compensation was performed using AbC total compensation beads (Thermo Fisher Scientific, cat. A10497) and ArC amine reactive compensation beads (Thermo Fisher Scientific, cat. A10346). Data was acquired on the FACSymphony A3 (BD Biosciences) cell analyzer system using the associated data acquisition software FACSDiva (v9.1; BD Biosciences). The gating strategies for immunophenotyping by flow cytometry are provided in Extended Data Figures 6-7.

#### IFN-gamma/Granzyme B ELISpot assay

The Human IFN-γ/Granzyme B Double-Color ELISPOT assay (Cellular Technology Limited (CTL), cat. hIFNgGzB-1M) was conducted according to manufacturer’s protocol. Briefly, the PVDF membranes were pre-wetted with 70% ethanol and coated with Human IFN-gamma/Granzyme B Capture Solution at 4°C overnight. Isolated patient PBMCs were thawed and recovered in Human T cell Medium (Gibco, cat. 0870112-DK) with 5% human serum (Valley Biomedical, cat. HP1022HI) overnight before being suspended in CTL-Test Medium. PBMCs were plated at a concentration ranging from 8×10^5^cells/well to 2×10^5^cells/well in a serial dilution. Cells were then stimulated with PepMix HCMVA pp65 peptides pool (JPT Peptide Technologies, cat. PM-PP65-2) at a final concentration of 0.2μg/well or with DMSO at a final concentration of 0.4μl/well as the vehicle control. T cell mitogen Phytohemagglutinin (PHA-P; InvivoGen, cat. inh-phap) was used as the positive control at a final concentration of 1μg/well while CTL-Test Medium alone served as the negative control. The plates were incubated at 37° in a CO_2_ incubator for 20 hours followed by the addition of anti-human IFN-gamma/ Granzyme B Detection Solution and further incubation at room temperature for 2 hours. Plates were washed and then incubated with Tertiary Solution at room temperature for 1 hour. Following washing, the plates were developed and spots were counted using the Cellular Technology Limited S6 Ultimate M2 Analyzer (service provided by CTL). The mean number of pp65 specific spot forming T cells from duplicate wells was adjusted to 2×10^5^ PBMCs.

#### TCR spectratyping

Sequencing of TCRβ VDJ rearrangements was performed using immunoSEQ Assay (Adaptive Biotechnologies, Seattle, WA). By means of an unbiased multiplex PCR, isolated genomic DNA was amplified and subsequently sequenced. The absolute number of TCRβ templates was determined and used for downstream analysis as previously reported.^41^ Cytomegalovirus (CMV)-associated TCRβ and corresponding complementarity-determining region 3 (CDR3) sequences previously determined^42^ were used to evaluate their presence in human patient samples using descriptive analysis. We also matched the significantly expanded TCRβ repertoire present, determined by differential abundance (Adaptive Biotechnologies), in our human patient samples to the pp65-associated TCRβ sequences curated by VDJdb database, including all scores^43^.

### Statistical Analysis, Rigor, and Reproducibility

Unless stated otherwise in the figure legend, independent group comparisons were analyzed using an unpaired Student’s t-test and ANOVA method. Pre-and-post groups’ difference and change over time in RNA-LPA ratios were evaluated using mixed effect models. Time-to-event data were analyzed using the Kaplan-Meier method along with the log-rank test. All p-values were reported as exact values, and the level of significance was denoted as follows: *p<0.05, **p<0.01, ***p<0.001, ****p<0.0001. Experimental results were repeated across model systems to validate results using different tumor models, and different species (mouse, canine, and human). Experimental data for human subjects were subject to third-party oversight and validation. Statistical software programs used for analysis were GraphPad Prism 9 and R 4.2, in a reproducible manner, using R markdown.

## Notes

### Clinical Trial

NCT04573140

### Funding Statement

This work was supported by federal awards W81XWH-17-1-0510; K08CA199224; R37CA251978; R01CA266857; R01FD007268 (FDA Office of Orphan Products Development); Florida Department of Health 20B11 (Bankhead Coley) and 20L07 (Live Like Bella) awards; and foundation grants from CureSearch (Catapult Award); The Rally Foundation; Hyundai Hope on Wheels (Hope-Scholar Award); St. Baldrick's; Chance for Life; Team Jack; Stop Children's Cancer; Danny's Dream; and The National Pediatric Cancer Foundation. Funding is also supported in part by the University of Florida Interdisciplinary Center for Biotechnology Research with funding provided by NIH Grant (1S10OD020026). J.G. acknowledges support R35GM146877 from NIGMS.

### Author Declarations

IRB of University of Florida gave ethical approval for this work

