## Extended Data Figures for "mRNA aggregates harness danger response for potent cancer immunotherapy"

### Extended Data Figure 1

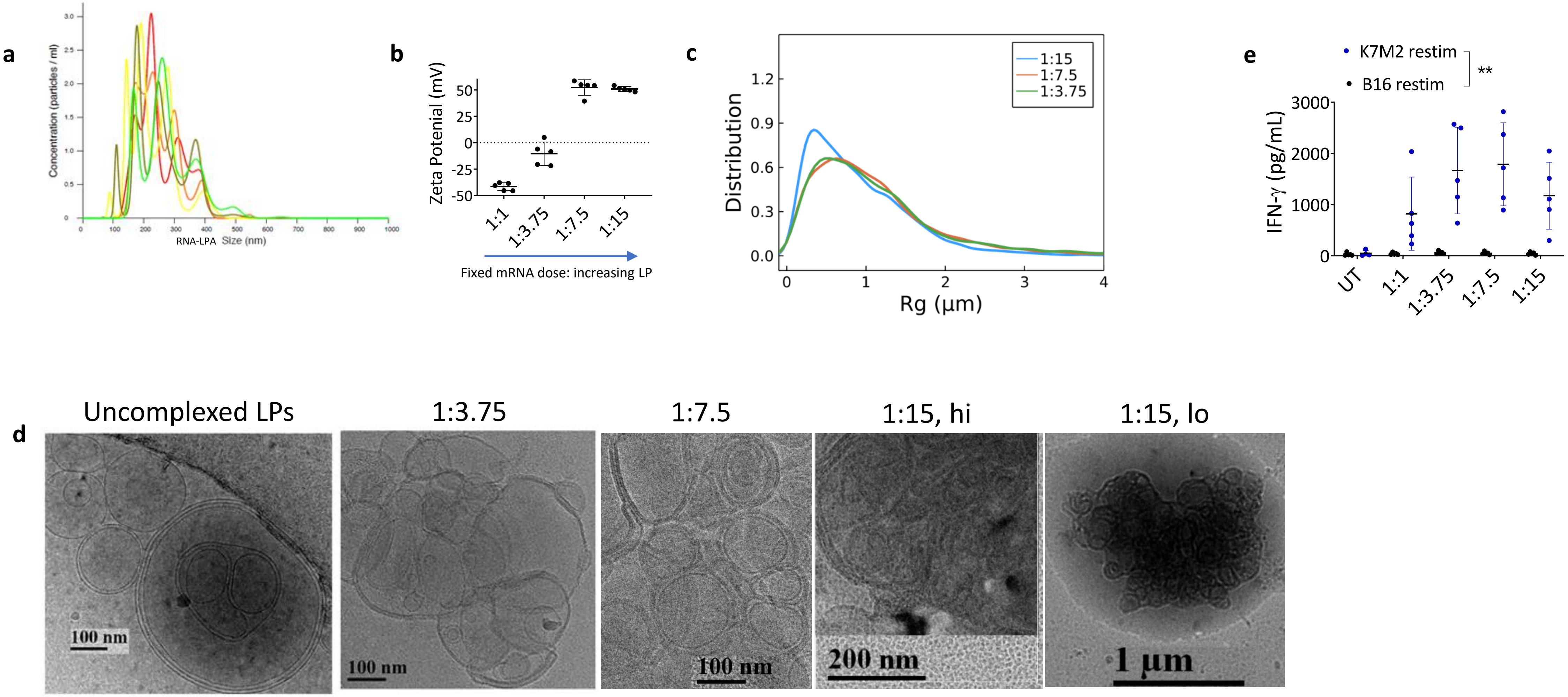

### Extended Data Figure 2

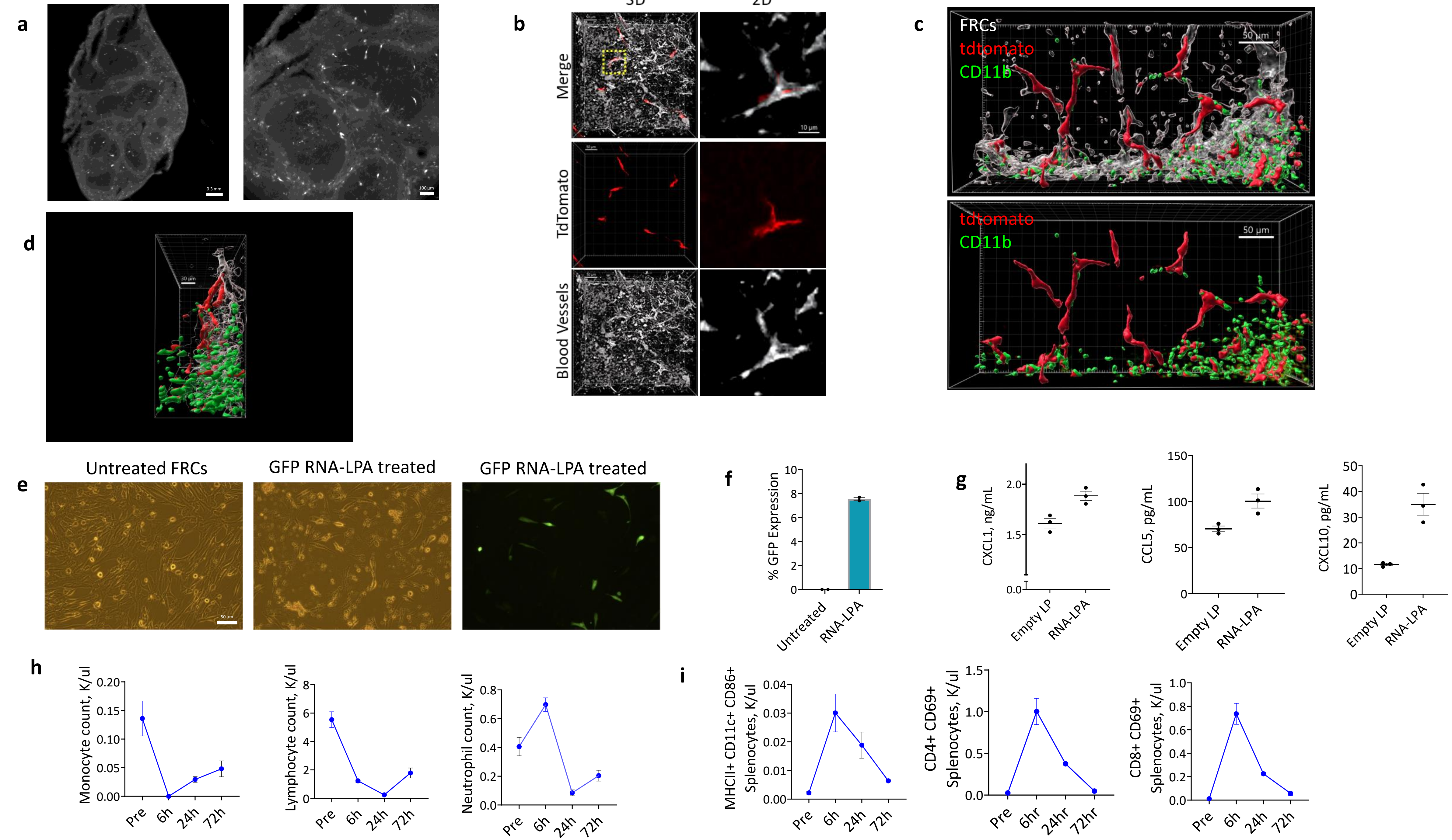

### Extended Data Figure 3

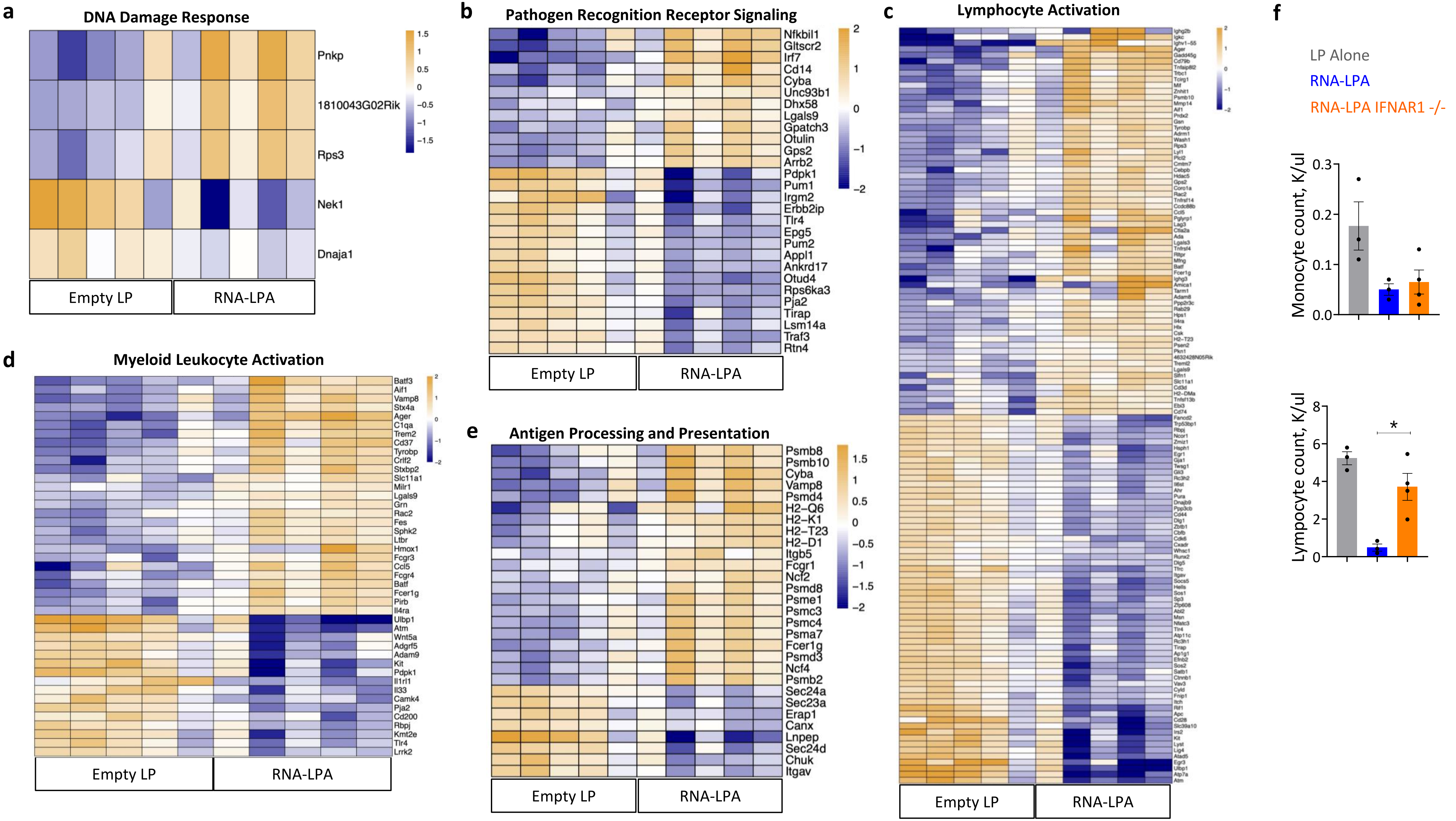

### Extended Data Figure 4

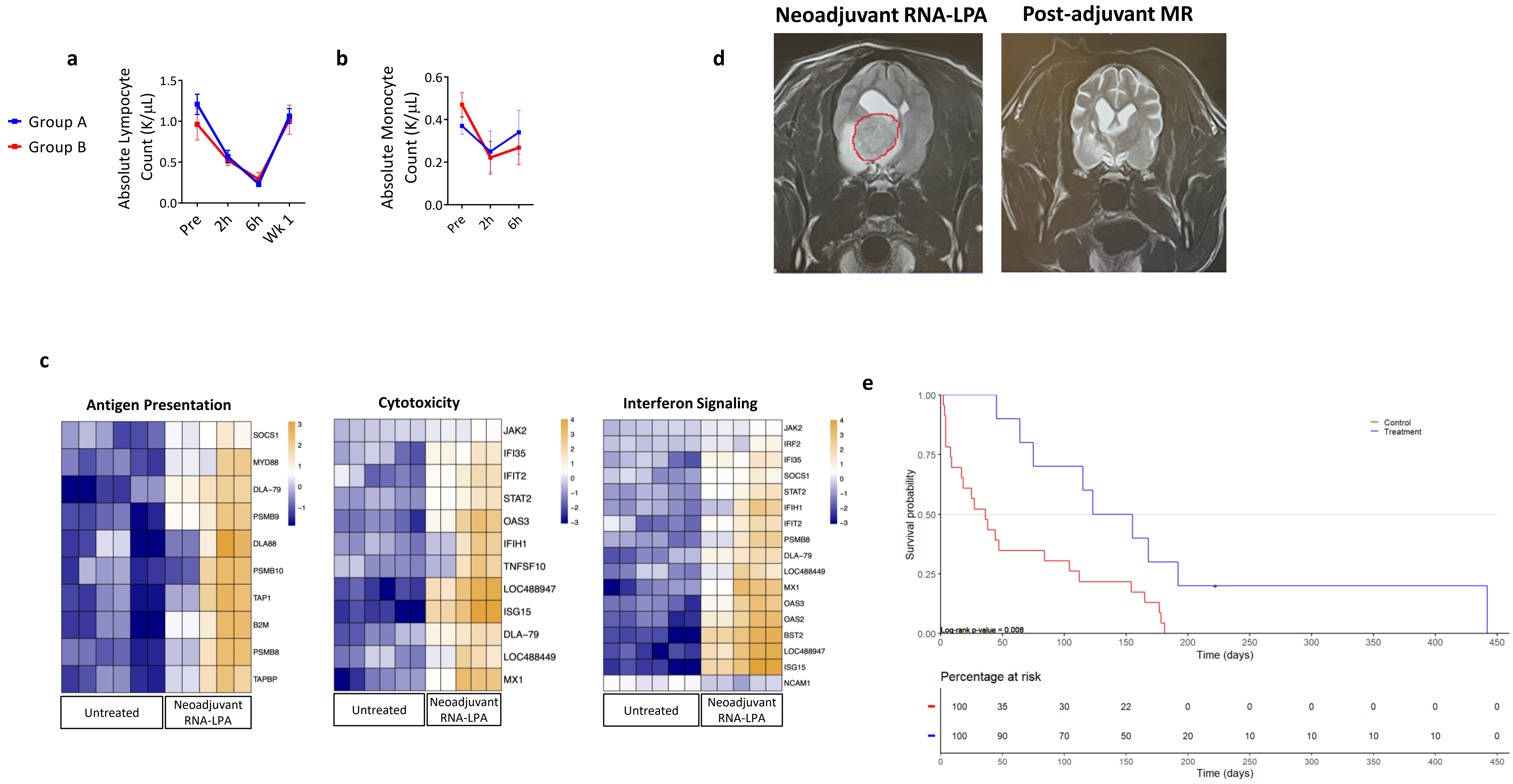

### Extended Data Figure 5

**a**

| Group | Total Number of Animals (Sex) | Subgroup (Termination) | Test Article | Route |
|---|---|---|---|---|
| 1 | 10 (5M/5F) | 1a (35±1 days) | Vehicle | i.v. |
| | 10 (5M/5F) | 1b (56±2 days) | | |
| | 10 (5M/5F) | 1c (112±3 days) | | |
| 2 | 10 (5M/5F) | 2a (35 ± 1 days) | LP | i.v. |
| | 10 (5M/5F) | 2b (56±2 days) | | |
| | 10 (5M/5F) | 2c (112±3 days) | | |
| 3 | 10 (5M/5F) | 3a (35±1 days) | RNA-LPA (low) | i.v. |
| | 10 (5M/5F) | 3b (56±2 days) | | |
| | 10 (5M/5F) | 3c (112±3 days) | | |
| 4 | 10 (5M/5F) | 4a (35 ± 1 days) | RNA-LPA (High) | i.v. |
| | 10 (5M/5F) | 4b (56±2 days) | | |
| | 10 (5M/5F) | 4c (112±3 days) | | |

**b**

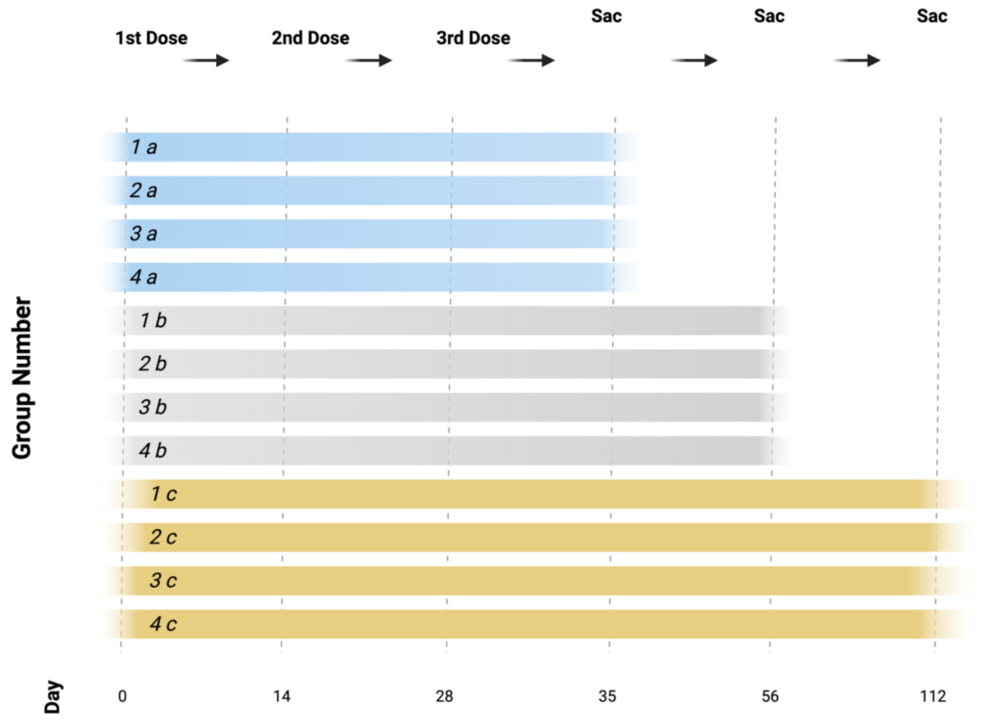


**c**

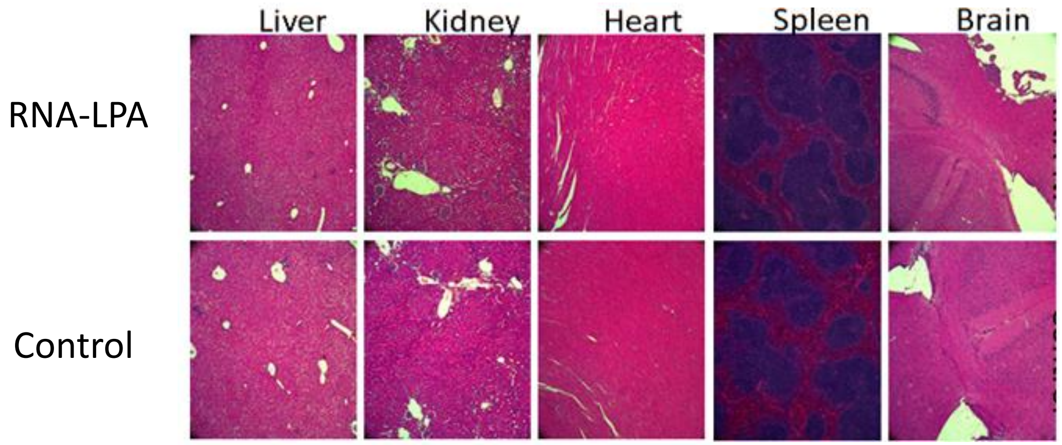

### Extended Data Figure 6

**a**

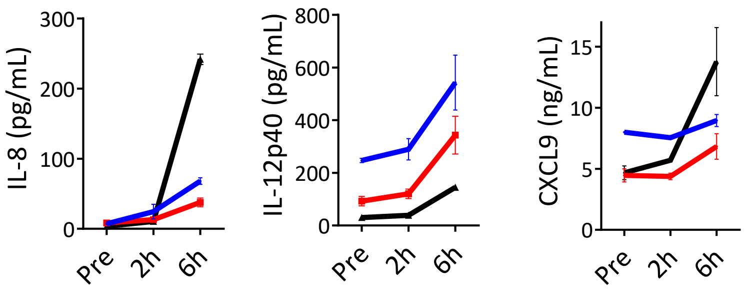


**b**

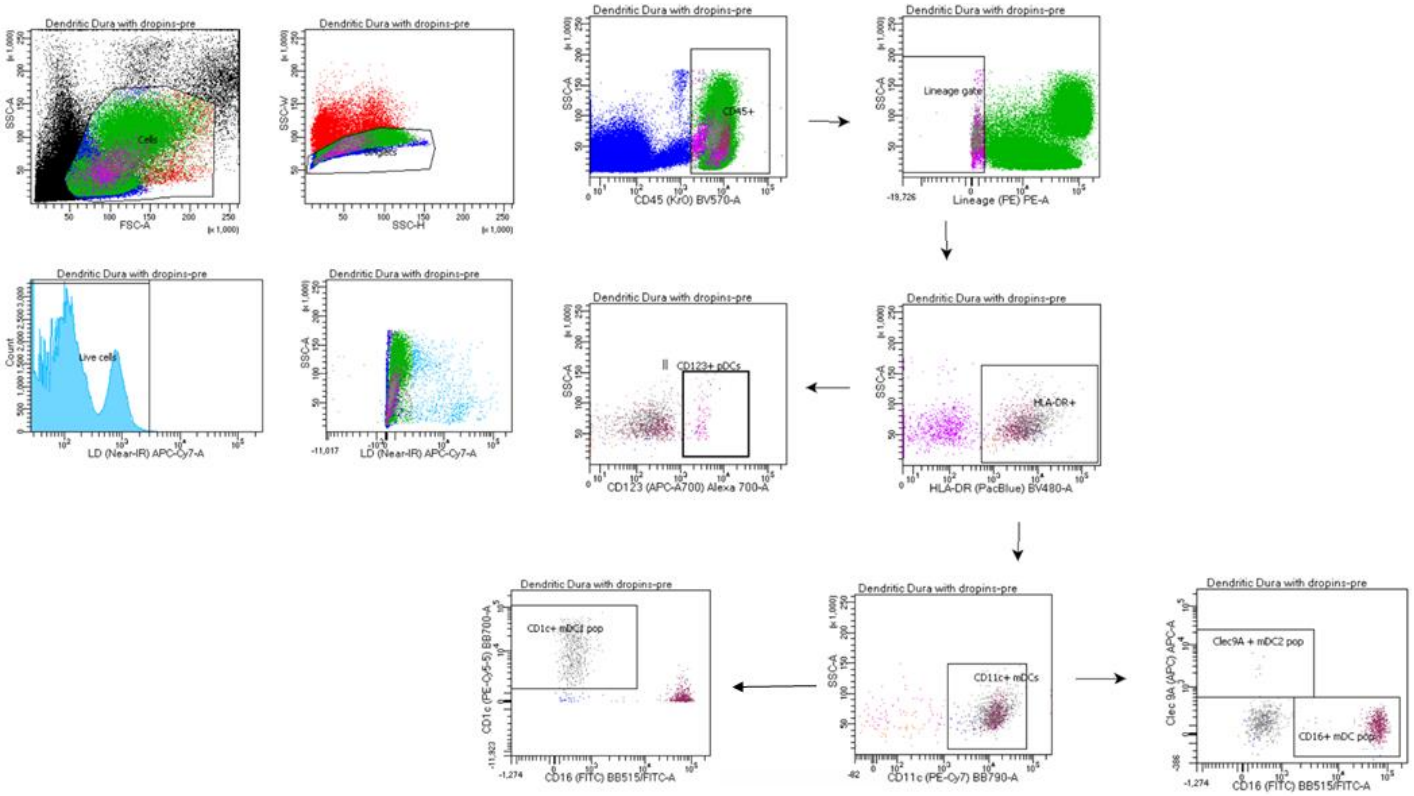

### Extended Data Figure 7

a

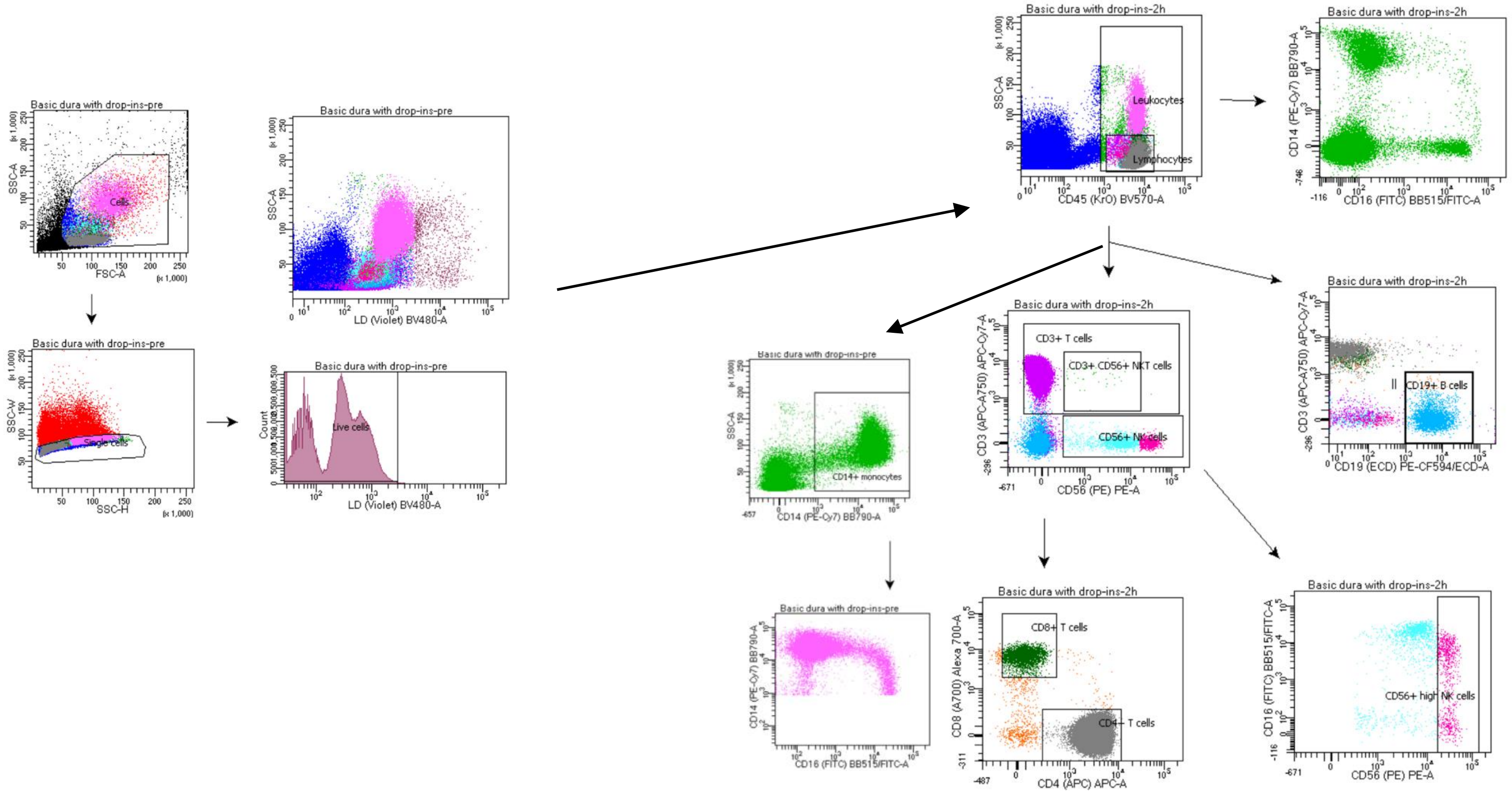


b

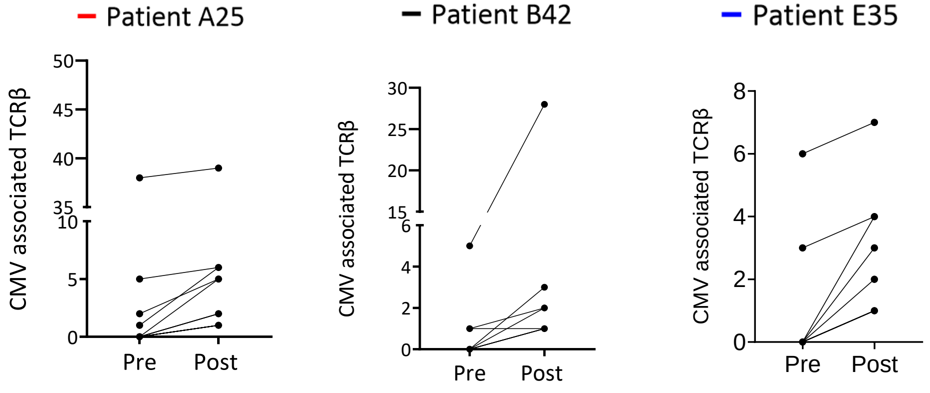

### Extended Data Figure 8

a

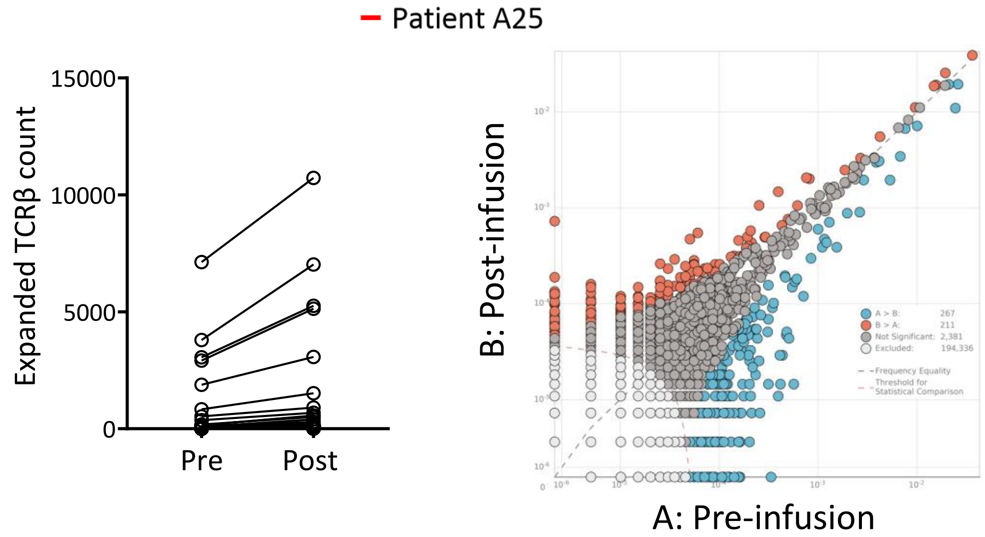


b

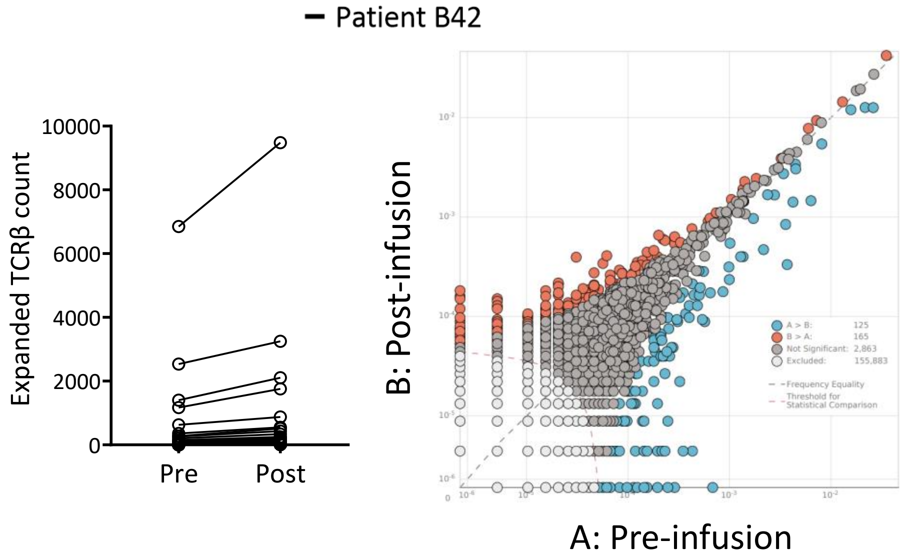


c

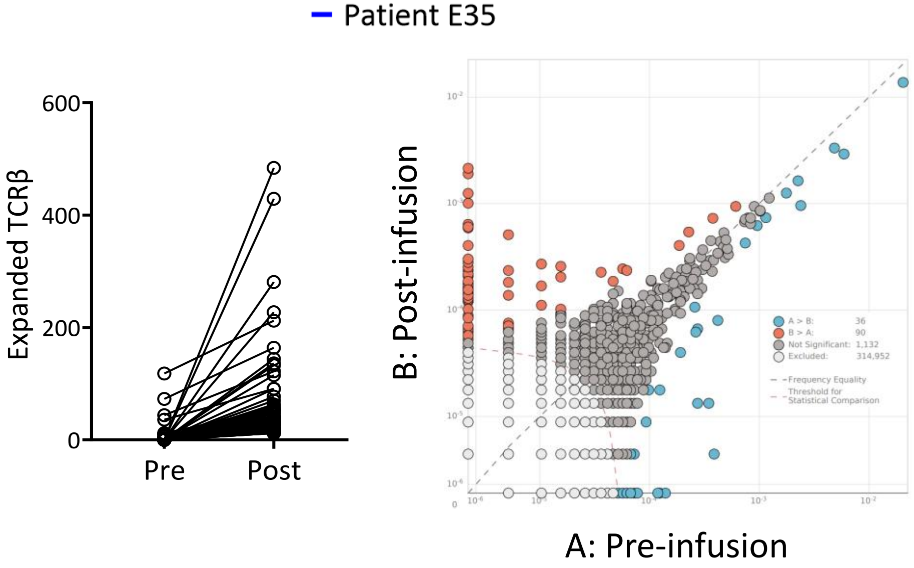
